# Immediate to longer-term neurophysiological impact of acute neural network disruption

**DOI:** 10.64898/2026.05.23.26353585

**Authors:** Zsuzsanna Kocsis, Ryan M. Calmus, Jithendar Kasa, Joel I. Berger, Ariane Rhone, Grant Brown, Carolina Deifelt-Streese, Mark Bowren, Peter N. Taylor, McCall E. Sarrett, Inyong Choi, Bob McMurray, Hiroto Kawasaki, Timothy D. Griffiths, Matthew A. Howard, Christopher I. Petkov

## Abstract

Despite substantial interest in how neural systems recover over time after acute neurological events, there is a dearth of longitudinal assessment from minutes to months surrounding these events. We compared rare intraoperative recordings in three patients, obtained immediately before and after anterior temporal lobe resection during a semantic prediction task, with longitudinal source-localized electroencephalography (EEG) obtained 2-6 weeks before, and 2 and 6-14 months after surgery. Relative to controls (*n* = 20), task performance showed sustained impairment in the two left-hemisphere patients and delayed impact in the right-hemisphere patient. Consistent with theories on ipsilateral and contralateral hemisphere compensation, all patients exhibited bilateral EEG alterations in speech responses and effective connectivity that did not fully recover to pre-operative levels. Direct dataset comparisons of intrinsic neurophysiological biomarkers associated with timescales of processing (*τ*_INT_) and excitatory-inhibitory balance (aperiodic slope, χ_SPEC_) showed a striking months-long reduction in rapid neural timescales of processing and gradually increasing aperiodic slope (putatively reflecting increased cortical inhibition or reduced excitation) specifically in the ipsilateral hemisphere of all three patients. Amidst these neurophysiological alterations, semantic task performance did not return to pre-operative levels. These rare longitudinal data advance a mechanistic framework to non-invasively evaluate neurophysiological impact over multiple timeframes in other patient cohorts.

**Significance statement:** We report rare longitudinal recordings obtained minutes to months before and after epilepsy surgery. Using state-of-the-art measures of neurophysiological biomarkers and network dynamics, we observed immediate disruption followed by consistent, longer-lasting alterations in neural excitability and temporal functionality.

## Introduction

Understanding the behavioral and neurobiological consequences of neurological events, how neural systems compensate, and the time course of recovery remain clinical and scientific research priorities^1,2^. Neurophysiological recordings with epilepsy patients provide an ideal opportunity to reference state-of-the-art non-invasive measures^2-4^ with intracranial recordings for single-subject inference in understanding effects that are variable immediately after surgery.

Several longitudinal fMRI studies imaged patients undertaking language^1,5-7^ or memory^2,8-13^ tasks before and after anterior temporal lobe resection to treat intractable temporal lobe epilepsy (TLE). The studies advanced theoretical frameworks on how neural systems compensate after neurological events. Bonelli and colleagues motivated a *functional adequacy* hypothesis, which posits that neural system compensation is driven by enhancement of task-related activity in remnant areas of the ipsilateral hemisphere, led by brain areas capable of compensating for memory or language impairment^12^. A not mutually exclusive *contralateral reserve* hypothesis has also found support, whereby homotopic areas in the contralateral hemisphere show task-related activity enhancement that can rescue mnemonic function, although language-related functions appear more resistant to recovery after left hemisphere surgery^14^. Recent longitudinal fMRI studies have broken into the realm of decade-long assessment, demonstrating that neural system alterations in patients can progress for a decade, exhibiting neither full return of pre-operative function, nor return to levels defined by controls^1,2^.

Precision site-specific neurophysiological recordings can be obtained in epilepsy patients during clinical monitoring via implanted intracranial electrodes. However, because intracranial electrodes are typically explanted before clinical resection, intracranial recordings are rarely obtained after surgical resection to treat intractable TLE^15^. We previously reported on fronto-temporal intracranial recordings obtained tens of minutes before and after left-hemisphere anterior temporal lobe (ATL) disconnection in two patients who participated in an intraoperative semantic expectancy task designed to engage the ATL^16^. The prior study obtained evidence in support of a modern diaschisis hypothesis, whereby remnant ipsilateral fronto-temporal areas demonstrated incomplete functional compensation. This was observed as alterations in neurophysiological responses to speech sounds and task-related effective connectivity that did not return to pre-operative levels in the minutes after resection^16^. It was not possible to evaluate the potential compensatory role of the contralateral hemisphere, or how the immediate neurophysiological impact of resection evolves over longer timeframes towards recovery.

Here, we aimed to substantially extend previous insights into the impact of surgery in the two left hemisphere patients^16^, supplemented with an additional dataset now available in a right hemisphere TLE resection patient. Longer-term assessment relied on source-localized scalp EEG recorded during the same task obtained 2-6 weeks pre-operatively, 2 months post-surgery, and 6-14 months post-surgery. We first evaluated source-localized EEG responses for site-specific reduction or enhancement of speech responses and effective connectivity in ipsilateral and contralateral hemisphere regions, focusing on auditory cortex and the inferior frontal gyrus (IFG) as candidate areas capable of compensating for the loss of the ATL in the semantic knowledge network^17^. We then directly compared the patients’ intracranial and EEG data using two prominent intrinsic neurophysiological biomarkers: intrinsic time constant, *τ*_INT_ ^18^, associated with neural timescales of processing; and spectral domain *1/f* aperiodic slope, χ_SPEC_, putatively reflecting excitatory-inhibitory balance^19^. As a longitudinal within-subjects study, the analyses focus on pre- and post-operative comparisons in the patients, referenced to healthy control EEG data (*n* = 20) whenever possible, and we only interpret effects that are significant and consistent in directionality in the patients.

We tested several hypotheses (Fig. 1A): 1) *Functional recovery*: We expect short-term functional impact^1,2,20^, but tested for a longitudinal return to pre-operative function. 2) *Classical diaschisis or only disruption hypotheses*: We tested for *reduction* (or *disruption* in variability) of neurophysiological responses and biomarkers in remnant brain areas^21^. 3) *Incomplete functional compensation*: The modern diaschisis hypothesis posits that site-specific enhancement of function reflects attempts at neural system compensation^16,22^, tested with the specificity of the functional adequacy and contralateral reserve hypotheses^12^.

**Figure 1.**
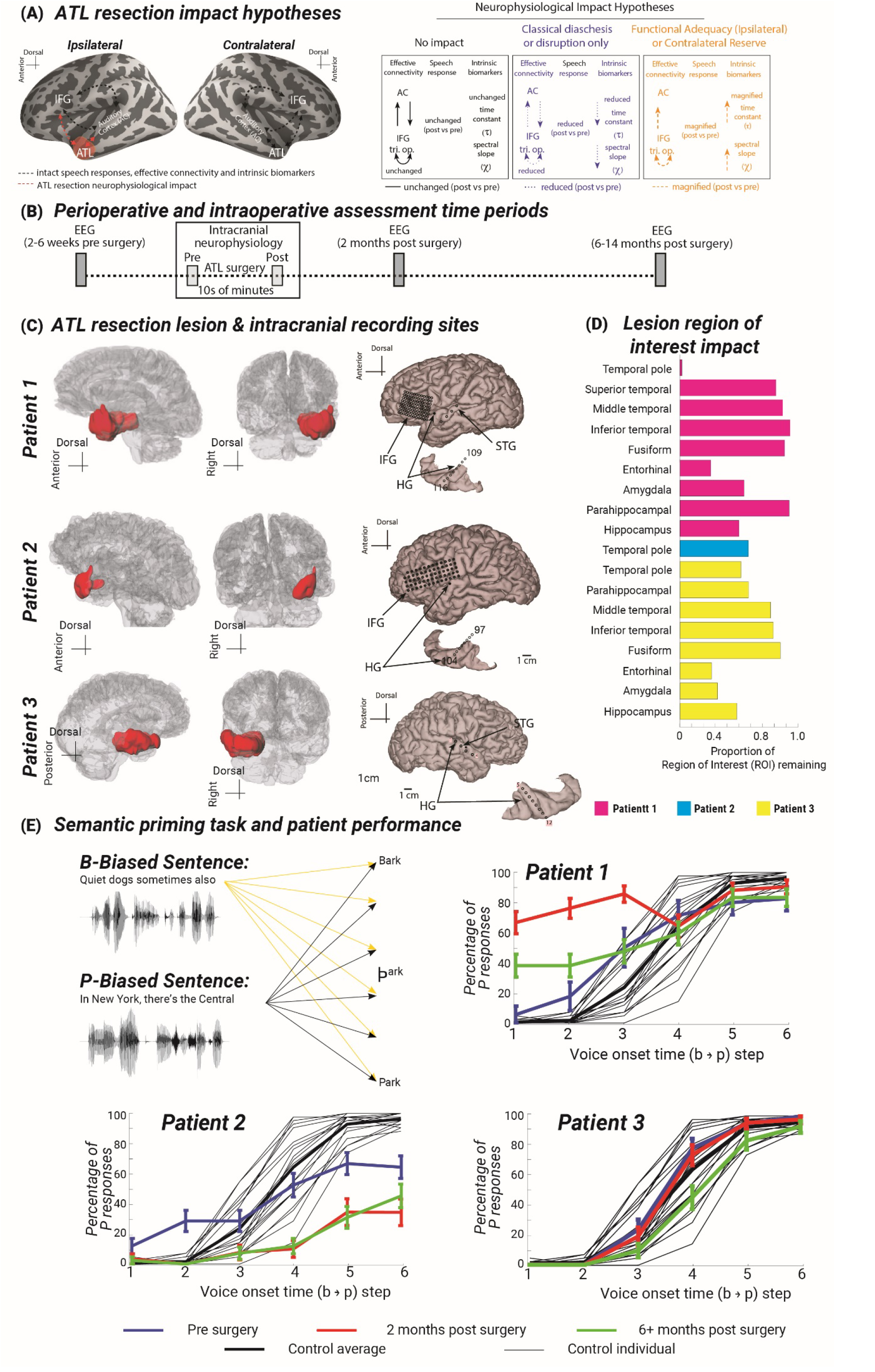
Brain areas and semantic expectancy task performance affected by the surgical procedure in the epilepsy patients. (A) Hypotheses regarding neurosurgical impact. (B) Timeline of perioperative (EEG) and intraoperative (intracranial recording) assessment time periods. (C) Post-operative T1-weighted MRI images showing the resected regions in red in each patient, including ATL and anterior medial temporal lobe resection in P1 and P3, but only the temporal pole in P2 to resect an epileptogenic cavernoma. Intracranial recording electrode locations are also shown per patient. (D) Proportion of each region of interest (ROI) remaining in each patient. (E) Depiction of the speech prediction paradigm and behavioral responses. Participants listened to /b/ or /p/ word biasing sentences with the final target word manipulated along the /b/ to /p/ voice onset time (VOT) steps. Responses shown as a function of the /b/ to /p/ VOT continuum and the percentage of /p/ responses given by the patients. The thin black lines show the control participant variability (*n* = 20), and the thick black line shows the average psychometric function of the controls. Blue lines show the average responses of the patients before the surgery, along with the standard error of the mean (SEM) across trials. The red lines show the patients’ average responses 2 months after their resection along with the standard error of mean (SEM) across trials and the green line 6+ months after surgery.

## Methods

For complete materials and methods, see the Supplementary Methods. The three patients in this study (P1: 61-65 year female, P2: 31-35 year male, P3: 35-40 year male) provided informed consent to participate under Institutional Review Board approval at the University of Iowa (IRB #200112047 and #201911155), consistent with the Declaration of Helsinki. Research participation was voluntary, could be withdrawn at any time, and did not affect clinical treatment. The patients had medically refractory epilepsy and underwent an awake craniotomy with language mapping requiring intracranial recordings in the operating room (Fig. 1C-D).

### Neurophysiological recordings and analysis

We previously described the intracranial recording approach^16^. Source-localized EEG signals were obtained before surgery, two months and six to fourteen months (P1: 6 months, P2: 14 months, P3: 8 months) post-operatively, while the participants performed the semantic expectancy task (Fig. 1E).

### Control participant EEG data

Source-localized EEG signals from twenty healthy control participants (average age: 19.26 years, SD = 1.8 years, 11 female) were obtained with the task for reference.

### Effective connectivity

Time-domain state-space Conditional Granger Causality (CGC) was computed on source-reconstructed time series, using the MVGC toolbox^23^ on per-trial epochs between -115 and 850 ms of the target word presentation onset.

### Intrinsic neurophysiological biomarkers: Time Constant (*τ*_INT_)

For the intracranial and EEG datasets we analyzed, respectively, the 1 to 1.5 sec period of silence between the behavioral task trials. The source-localized EEG signal was extracted from the following regions-of-interest (ROIs)—Heschl’s Gyrus (HG), Superior Temporal Gyrus (STG), IFG pars opercularis (IFGop), and IFG pars triangularis (IFGtri)—across both hemispheres (ipsilateral and contralateral to the site of resection) across the three timepoints. For each trial, the normalized autocorrelation function (ACF) was fit with an exponential decay model:

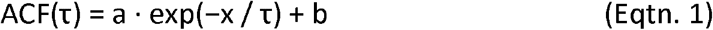

where *a* is the amplitude, *b* is a baseline offset, *x* is the lag in samples, and *τ* is the time constant in milliseconds.

### Spectral Domain Aperiodic Slope (χ_SPEC_)

Spectral parameterization was performed using FOOOF (Fitting Oscillations & One Over F^24^). The aperiodic (1/*f*) component was modelled in *fixed* mode (no spectral knee), with the exponent representing the spectral slope in log frequency.

### Statistical Analyses

For the brain-wide EEG signal (Fig. 2), linear mixed-effects (LME) model was evaluated:

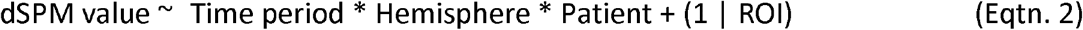

**Figure 2.**
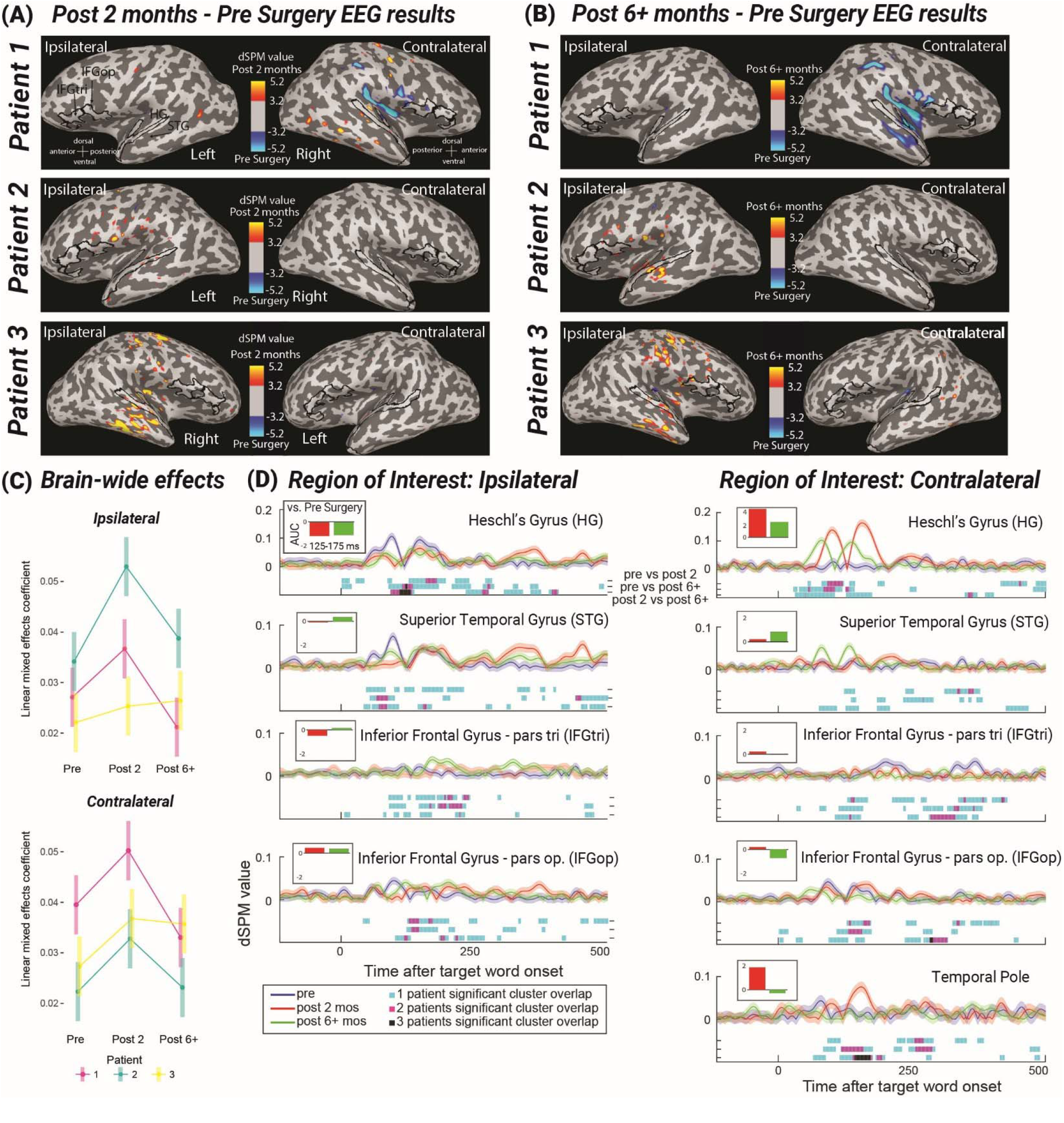
Source-localized EEG signal results for each patient. (A and B) Source-localized activation (dSPM values^30^) differences between the pre-surgical time point and two months (A) or 6+ months (B) after the surgery for each patient at ∼160 ms after target word onset, avoiding the non-specific initial sound driven response (see Suppl. Methods). Blue areas signify activation that was significantly larger at the pre-surgical timepoint (i.e., post-operative reduction); red areas indicate activation that was larger at the two or 6+ months post-surgical timepoint (i.e., post-operative enhancement). (C) Brain-wide ROI dSPM activation summary across the ipsilateral and contralateral hemispheres, shown by patient (see Suppl. Methods for the LME modeling approach). (D) Source-localized EEG activation (dSPM) of regions of interest averaged over patients. Activation is compared between pre-surgery vs. 2 months post-surgery, and 6+ months post-surgery, plotted together with the standard error of the mean (SEM) between trials. Blue line plots show the pre-surgical activation; red lines denote 2 months after the surgery, and green denotes 6+ months after the surgery. The lines under the plots show the temporally statistically significant cluster test (*p* < 0.05) differences with one, two, or three patients’ significant periods overlapping, compared between each time point (first row: pre-vs 2 months post-, second row: pre vs 6+ months post-, third row: 2 vs 6+ months post-operatively). In the bar graph subpanels for each ROI, we show the EEG activity response area under the curve (AUC) contrasted between the pre-resection and post-resection time points in the 125 to 175 ms time window after target word onset.

For ROI analyses, significance testing was conducted with cluster permutation to identify time periods of significant clusters overlap in the patients (*p* < 0.05, length > 5 ms). Pairwise CGC for every edge in each patient were thresholded to include 10% of the strongest edges log Granger’s *F* differences. Combined CGC graphs were masked by intersecting edges of identical sign across all patients, reflecting consistent directionality of effects.

Statistical inference for *τ*_INT_ and χ_SPEC_ was performed using LME with dependent variables: τ (ms) or aperiodic (1/*f*) exponent, including fixed effects of Hemisphere, ROI, and Time period (pre, 2 & 6+ months post):

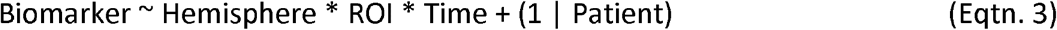

To adjust for the large biomarker sample sizes (per trial, region, hemisphere and patient), we applied a stringent Bonferroni correction with a corrected *p* < 0.001 acceptance threshold. For tests of *τ*_INT_ distribution differences, we evaluated Bowley skewness, *B* = (Q3 + Q1 − 2·Q2) / (Q3 − Q1), where Q is the quantile level for each distribution. Suppl. Tables 3-7 show complete statistical testing outputs.

## Results

All three patients showed significant semantic expectancy task performance effects over time. The two left hemisphere patients’ performance was significantly degraded pre-versus post-operatively at both 2 and 6+ months (P1 and P2, all *p* < 0.001). The right hemisphere resection patient’s performance was unaffected pre-versus 2 months but significantly differed in the pre-versus 6+ and 2 versus 6+ month assessment periods (*p* < 0.001; Fig. 1E, Suppl. Table 3). All patients performed well on the control tasks (e.g., no lexical competition; Suppl. Table 3).

### Source-localized EEG and effective connectivity

The whole brain source-localized EEG responses to the target words showed largely ipsilateral hemisphere enhancement to the target word in all patients (red colormap in Fig. 2A-B). These brain-wide effects were evaluated with a Linear Mixed Effects model (Fig. 2C), showing significant post-operative enhancement in the target word response in all patients pre-versus 2 months post-operatively (all *p* < 0.01, except P3 in the ipsilateral hemisphere; Suppl. Table 4). Figure 2D shows the post-operative alterations in target word responses within the putative compensatory semantic knowledge network, revealing similar temporal alterations of low-frequency responses to the target word in the ipsilateral hemisphere as reported previously for P1 and P2^16^ and shown for P3 in Suppl. Fig. 3.

We evaluated directed effective connectivity during the target word response using the CGC analyses. Figure 3 shows the alterations in effective connectivity across time. At both 2 and 6+ months post-versus pre-surgery, we observe *decreases* in *intra-hemispheric* effective connectivity (blue edges; at 2 months: total decreased intra-hemispheric versus total edges, 8/16 edges; 6+ months: 14/18 edges), including in interaction with the remnant contralateral temporal pole. Comparing 6+ to 2 months post-operatively, we observe marked *increases* in connectivity between these two post-operative periods (red edges), with nearly all consistent *increases* in effective connectivity during this period *inter-hemispheric* in nature (7 of 8 edges).

**Figure 3.**
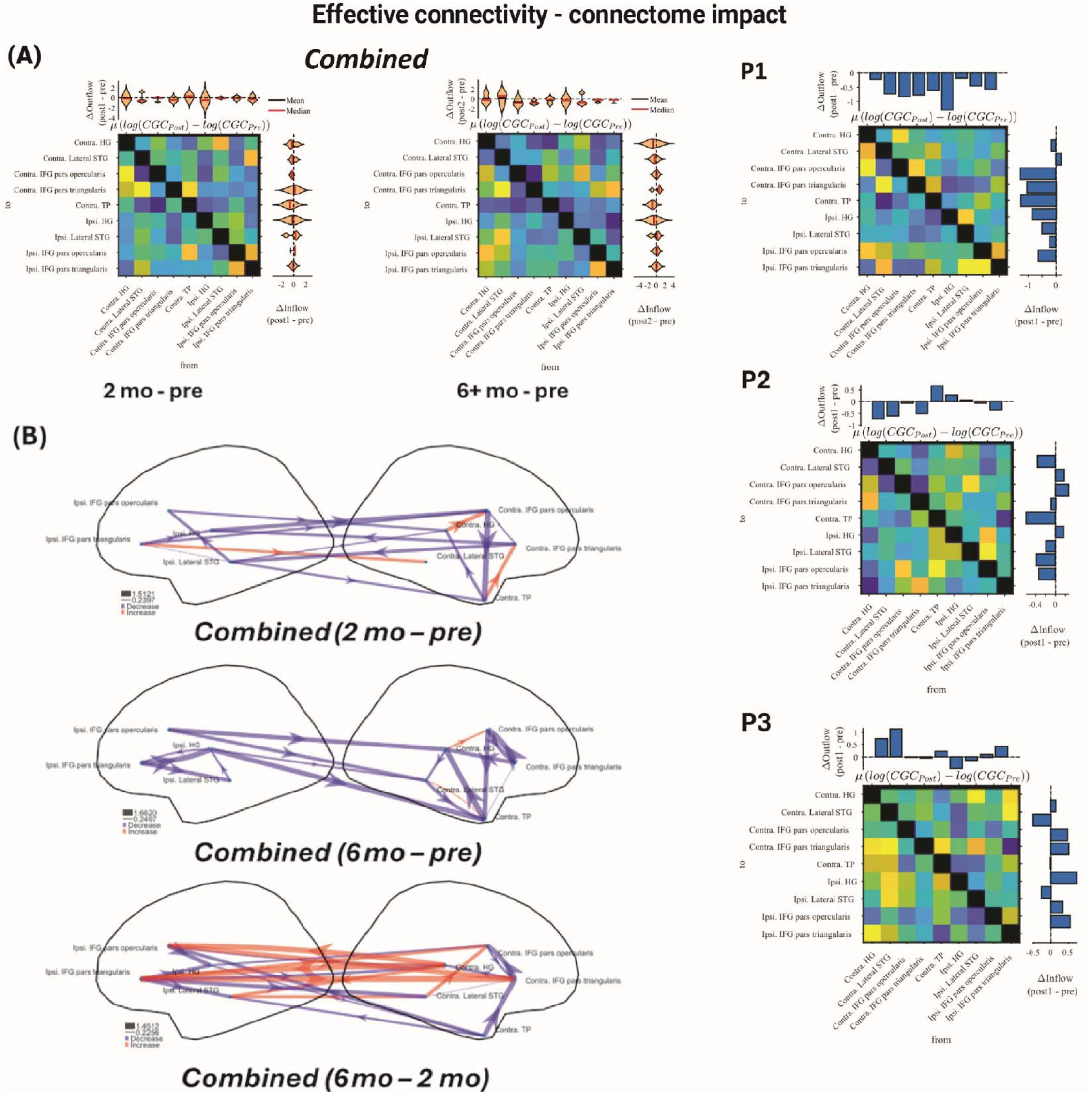
Effective connectivity results via state-space Conditional Granger Causality (CGC). (A) Mean post-minus-pre log-CGC values across the 3 patients, computed on log-transformed Granger’s *F* (Methods). Changes in outflow/inflow from/to each node were computed on these values summed across rows/columns, respectively. Per-patient 2-month post-versus pre-surgery matrices are also shown. (B) Graphs of mean log-CGC changes are masked to retain only those changes matched in directionality (sign) across *all* 3 patients (blue edges: decreases; red: increases; edge width proportional to magnitude of CGC change).

### Intrinsic neurophysiological biomarkers

Our first intrinsic neurophysiological biomarker is the slope of the spectral aperiodic component χ_SPEC_ (Fig. 4A)^19^. Intraoperatively, the spectral slope was steeper in the patients relative to the EEG data in controls, which may reflect reduction in excitability driven by anesthetic management (Fig. 4C; all *p* < 0.001 corrected; Suppl. Table 5). Divergent post-resection spectral slope effects were observed intraoperatively post-versus pre-surgery (P1 decreased; P2 not significant; P3 increased; Suppl. Table 5). By contrast, pre-operative EEG aperiodic slope was within the range of the control participants, and there was a months-long increase in the aperiodic slope in the two post-operative assessment periods in all three patients primarily in the ipsilateral hemisphere (Fig. 4D; *p* < 0.001 corrected, Suppl. Table 5; by-patient plots in Suppl. Fig. 6).

**Figure 4.**
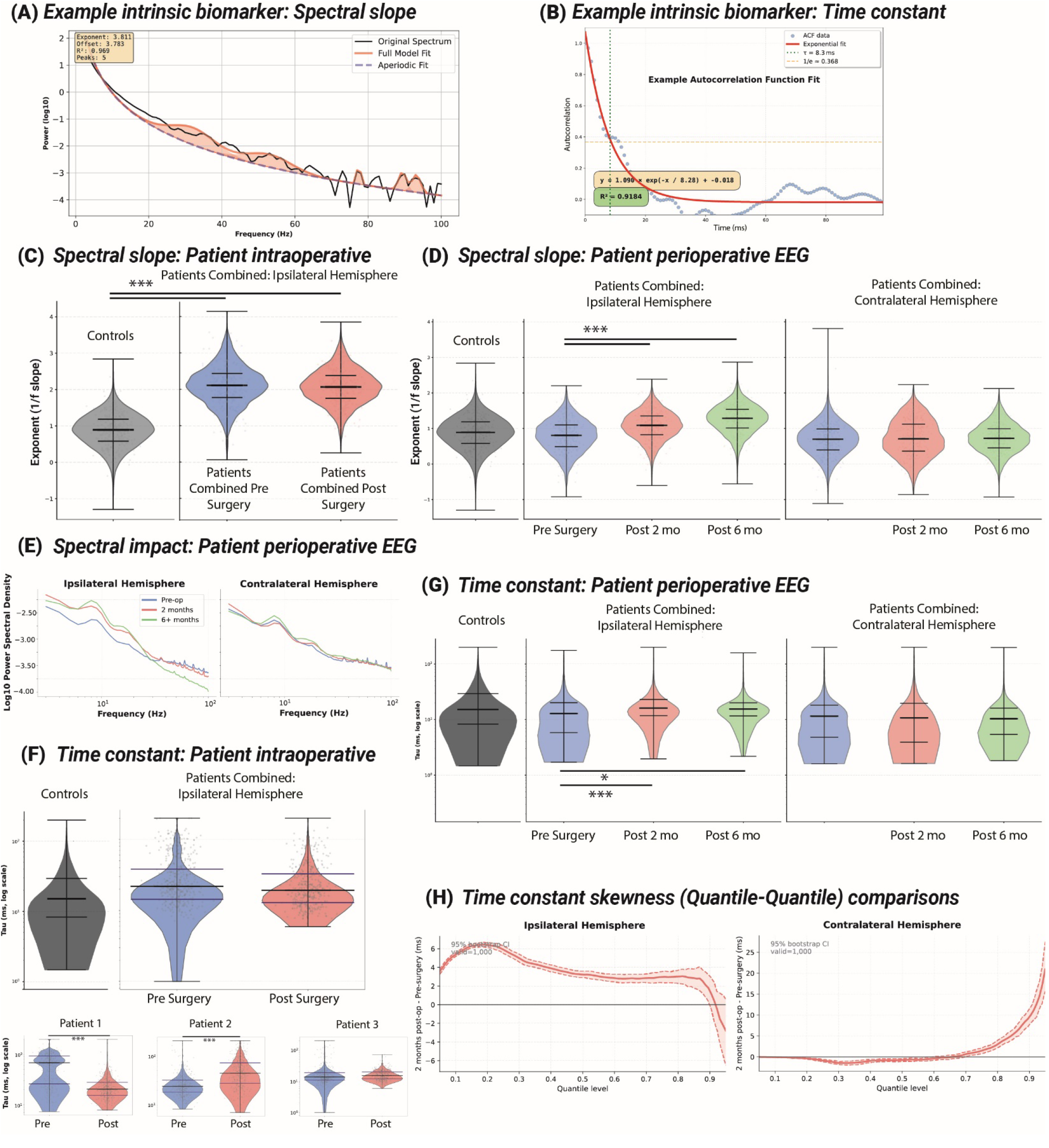
Resection impact on two intrinsic neurophysiological biomarkers. (A) Example aperiodic (1/f) spectral slope (χ_SPEC_) exponent calculation with FOOOF^24^, related to panels C-E. (B) Example autocorrelation function fit to extract the intrinsic time constant (*τ*_INT_), related to panels F-H. Both were fit on 1 or 1.5 s samples during silent periods between the semantic task testing trials. (C-D) Violin plots of the distributions of spectral slope exponent values (blue: pre-surgery; red: 2 months post-surgery; green: 6-14 months post-surgery), referenced to control participant EEG distributions (gray). (C) Intraoperative patient recording χ_SPEC_ results. (D) Longitudinal EEG χ_SPEC_ results. For C-D, *** indicates *p* < 0.001 corrected central tendency differences (Suppl. Table 5). (E) Grand average spectral power separated by hemisphere, showing results consistent with increased spectral slope (compare with D) post-operatively in the ipsilateral hemisphere, seen as a combination of increased low frequency (< 20 Hz) and decreased higher frequency power (> 20 Hz). (F-G) Time constant distributions intraoperatively (F) and perioperatively with EEG (G). For F and G, *** indicates *p* < 0.001, ** *p* < 0.01 and * *p* < 0.05 corrected central tendency differences (Suppl. Table 6). Distribution skewness test results shown in Suppl. Table 7. (H) Time constant skewness Quantile-Quantile (Q-Q) comparisons.

We also evaluated the time-domain *τ*_INT_ associated with neuronal timescales of processing^18^ (i.e., time constant of the autocorrelation decay function; Fig. 4B). Intraoperatively, the distribution of observed time constants was within that of controls (Fig. 4F; *p* > 0.05 corrected; Suppl. Table 6). Intraoperatively, as noted above in the immediate differential impact on the aperiodic slope (Suppl. Table 5), post-resection time constant effects differed (i.e., increased in P1, decreased in P2, no change in P3; Fig. 3F; all *p* < 0.05 corrected, Suppl. Table 6). In the months-timeframe perioperatively, EEG-derived time constant was within the distribution of control participants (Suppl. Table 6). However, post-operatively there was a substantial loss of short time constants, seen as narrower distribution—reduced lower quartile—in the ipsilateral hemisphere (Fig. 4G-H; Bowley skewness test, *p* < 0.05, Suppl. Table 7; Suppl. Fig. 8), largely affecting auditory cortex *τ*_INT_ values (Suppl. Fig. 7).

## Discussion

We report an initial longitudinal comparison of the minutes-to months-long neurophysiological impact of temporal lobe resection in three adult epilepsy patients. Both left hemisphere patients demonstrated sustained impairment in performance on the semantic prediction task, with a significant delayed impact on task performance in the right hemisphere patient (Fig. 1E). Intrinsic neurophysiological biomarker alterations showed a gradual increase in the aperiodic slope in the ipsilateral hemisphere and a striking reduction in short time constants, consistently in all patients (Fig. 4). Neither behavioral performance on the semantic expectancy task, nor the observed neurophysiological alterations completely returned to pre-operative levels, challenging the complete compensation hypothesis within the first year following resection (Fig. 1A). Outside of the operating room, the results also ruled out the classical diaschisis hypothesis (i.e., purely disruptive effects), supporting instead the ipsilateral functional adequacy and contralateral reserve hypotheses, both of which posit enhanced activity in remnant brain areas reflect neural system attempts at compensation for months following neural network disruption^12^.

Longitudinal whole-brain EEG analysis, comparing pre-operative to early post-operative (2 months) levels, showed ipsilateral and contralateral hemisphere enhancement post-operatively in response to the target word during the semantic expectancy task (Fig. 2C). This presented as combinations of site-specific time varying neurophysiological enhancement or reduction in target word responses, including auditory cortex and inferior frontal gyrus, candidate areas within the semantic knowledge network capable of compensation for loss of ATL function^17,25^. Similar alterations in the intracranial responses to the target word in the ipsilateral hemisphere were previously reported in P1 and P2^16^, and P3 here.

The intracranial recordings reflect the immediate impact of the resection, now extending up to a year after surgery in reference to the source-localized EEG data. The EEG results are generally consistent with several longitudinal fMRI studies following TLE resection in epilepsy patients using language-related tasks (such as verbal fluency and verb generation tasks), showing enhanced fronto-temporal activation in both hemispheres following resection^1,5,7^. Our effective connectivity results (using directional state-space CGC) further informed these observations, demonstrating strong post-operative decrease in interhemispheric effective connectivity relative to pre-operative levels. Interestingly, we also observed potential attempts at compensation that manifested as *enhancement* of inter-hemispheric interconnectivity between 2 and 6+ months post-surgery (Fig. 3B), within an otherwise deeply post-surgically reduced intra-hemispheric connectivity between the network nodes and the remnant contralateral temporal pole.

Intrinsic neurophysiological biomarkers can be efficiently derived from ‘resting-state’ signals, including during silent periods between task trials. Our first biomarker of interest was the 1/f exponent of the slope of the aperiodic spectral profile (χ_SPEC_ ^24^). The patients’ aperiodic slope was elevated relative to controls in the intraoperative setting, possibly because of the sedatives surrounding the awake periods in the patients, and the aperiodic slope was altered in different directions in each patient after surgery. By contrast, the EEG aperiodic slope was within the distribution of controls pre-operatively but increased in the 2 and 6+ month timepoints in all patients in the ipsilateral hemisphere, where a steeper aperiodic slope may be associated with greater neural inhibition or less excitation^19^.

The second biomarker of interest was the intrinsic time constant, *τ*_INT_, quantifying the reduction in self-similarity of the neurophysiological signal over time and associated with neural processing timescales^18^. While the patients’ pre-operative time constant distribution was within the range of controls, the resection yielded heterogeneous changes across patients after surgery, paralleling the results of our aperiodic slope analysis. Strikingly, post-operatively the distribution of time constants (*τ*_INT_) revealed a significant reduction in shorter time constants in the ipsilateral hemisphere (Fig. 4D), primarily affecting auditory cortex (Suppl. Fig. 7). We interpret this finding as a consequence of the loss of the ATL, specifically affecting rapid temporal processing functions in auditory cortex. Our observation is generally consistent with reporting that stroke can lengthen intrinsic time constants, consistent with slowed temporal dynamics^26^.

The key limitation of this study is its small sample size, which reflects the inherent rarity of consistently obtaining awake pre- and post-operative intracranial and longitudinal EEG recordings in general, and especially from patients performing language semantics tasks^14,27^. Epilepsy patients increasingly have access to a widening array of treatment options not requiring craniotomy^28^. Therefore, data like these are likely to become even more difficult to obtain. Nonetheless, analyses of the efficacy and impact of other epilepsy treatments will benefit from longitudinal neurophysiological assessment and benchmarking against the neurophysiological alterations shown here and the seizure-freedom levels of TLE resection^28^. For instance, thermal ablation of the medial temporal lobe using MRI-guided Laser Interstitial Thermal Therapy has been suggested to minimally impact cognition and language functions, a claim that is ripe for empirical testing^29^. In summary, this study provides novel mechanistic insights on the impact of and path towards recovery from surgical resection across timeframes from minutes to months, advancing a paradigm for neurophysiological assessment noninvasively, and in reference to intracranial recordings whenever possible^1,2^.

## Data Availability

All data produced in the present work are contained in the manuscript

## Acknowledgements

We thank the patients who took part in this research. We thank Athina Tzovara for the useful discussion. Funding support by National Science Foundation (SBE-2342847) and National Institutes of Health (U01NS13979575; S10OD025025; S10OD030220; S10RR028821; R01DC004290).

## Author contributions

⍰ **Conceptualization:** ZK, CIP, TDG & MAH

⍰ **Methodology:** ZK, RC, CIP, PNT, BM, IC, JK, AR

⍰ **Data Collection:** ZK, CD-S, AR, JB, HK, MES, BM

⍰ **Analysis:** ZK, RC, JB

⍰ **Writing - Original Draft:** ZK, CIP

⍰ **Writing - Review & Editing:** CIP, ZK, RC, AR, GB, JB, CD-S, PNT, IC, BM, MES, HK, MB, TDG, MAH

⍰ **Project administration:** CIP, MAH

## Data and code availability

The data and code will be made available with manuscript publication on the Open Science Framework: https://osf.io/arqp8/ under the Kocsis_et_al_Longitudinal_ATL folder.

