## Supplementary Materials for "Immediate to longer-term neurophysiological impact of acute neural network disruption"

### Supplementary Methods

Here we report the complete materials and methods associated with the manuscript. There may be some redundancy with the abbreviated version in the manuscript.

#### Materials and methods

The three patients within this study (P1: 61-65 year female, P2: 31-35 year male, P3: 35-40 year male) provided informed consent to participate under Institutional Review Board approval at the University of Iowa (IRB #200112047 and #201911155). Research participation was voluntary, could be withdrawn at any time, and did not affect clinical treatment.

Left hemisphere dominance in all three patients was established with a Wada test. The patients had medically refractory epilepsy and underwent an awake craniotomy with language mapping requiring intracranial recording electrode monitoring in the operating room. The resection of the seizure onset zone required left (P1 and P2) and right (P3) anterior temporal lobe (ATL) resection. P1 and P3 had a two-step surgical procedure, starting with ATL disconnection to access and then resect the deeper epileptogenic foci in the medial temporal lobe (MTL). For P2, the procedure only required ATL resection of the epileptogenic cavernoma located in the temporal pole. The post-operative intracranial recordings were obtained after the first step in each procedure, and the EEG recordings reflect electrophysiological responses within the combination of the resected tissue (Fig. 1C-D).

#### Neuropsychological assessment

The three patients were evaluated using a standard neuropsychological battery, as reported previously^1^. The list of tests administered and results are reported in Supplementary Table 1. In comparing the patients' pre-operative performance to population norms, *z*-scores were calculated using age- and education-matched means and standard deviations. To assess change in performance following ATL resection, we used the reliable change index (RCI). The index accounts for test-retest reliability to predict change over time^2^. RCI scores of ±1.96 are considered significant alterations between timepoints.

#### Semantic predictions task

We relied on an established semantic expectancy task designed to engage the ATL and fronto-temporal semantic knowledge network^1,3^. The task involves listening to sentences establishing an expectation in the perception of the final target word, which began with either a /b/ or /p/ phoneme (Fig. 1 and Suppl. Fig. 1). The target word was manipulated along a voice onset time (VOT) continuum (0 ms to 40 ms in 6 equally spaced steps) to alter perception of /b/ as in ‘beach’ (short VOTs) to /p/ as in ‘peach’ (long VOTs). Both endpoints were real words, creating naturalistic conditions where the biasing sentence and the target word either matched expectation established by the sentence preceding the target word or were incongruent. Sentence primes were counterbalanced with target words from each step of the VOT continuum^1,3^. During the task, 7 different word pairs were used (bill/pill, bad/pad, bath/path, back/pack, bark/park, bowl/pole, beach/peach), amounting to 504 experimental trials. We also included two types of control tasks: 126 filler trials that fully predicted the target word (so that participants expected a majority of trials to follow their expectations) and 63 catch trials where the target word was presented visually on a screen along with another written word to choose from (to ensure participants were attending to the semantic content of the sentence). These could either be fully predictive sentences, as in the case of the filler trials, or fully congruent sentences taken from the main experiment. The participants were asked to press a button after each sentence to indicate what they heard (/b/ or /p/ for experimental trials) or the word that best fit the sentence in the case of the catch trials. The number of trials the patients completed is shown in Suppl. Table 2.

For behavioral hypothesis testing, a linear mixed-effects model (LME) was fit for each patient, predicting /p/ responses from scaled VOTs (1-6 levels), Bias (2 levels: /b/ or /p/), and Time period of assessment (3 levels: pre-, 2 months post-, and 6+ months post-surgery) with random intercepts for word pair. Estimated marginal means for Time were obtained from the model, and Bonferroni correction was used for pairwise comparisons. The model in (Eqtn. 1) was fit using the *lme4* package in *R* with the following *lmer()* notation^4^:

/p/ responses ~ Time period + Bias + VOT + (1 | Word pair)	(Eqtn. 1)

#### Neurophysiological recordings and analysis

We previously described the intracranial recording approach^1^. High-density channel scalp EEG recordings were obtained before surgery, two months post-operatively, and six to fourteen months (P1: 6 months, P2: 14 months, P3: 8 months) post-operatively, while the participants performed the speech prediction task. On each of the assessment days, we obtained T1-weighted structural MRI scans (32-channel head coil, TR = 8.5, TE = 3.3, 1x1x0.8 mm voxel resolution) with a 3T Discovery MR750w scanner (General Electric, Wisconsin, USA), used for source localization. For the EEG recordings, a 128-channel Biosemi Active2 electrode system was used. Data were recorded at 2048 Hz, and electrodes were placed according to the international 10-20 system and referenced online to the Common Mode Sense (CMS) active electrode, placed to the left of the Pz electrode. Fiducial markers (nasion, left periauricular, and right periauricular) and electrode position were digitized using the Polhemus Patriot 6DOF motion tracking system and Cortech Locator software. EEG source localization was performed with MNE-Python software^5^. Data were downsampled to 1000 Hz and filtered offline between 3 and 30 Hz. Before source localization, bad channels were identified and dropped from further analysis (Suppl. Table 1). Source estimates were quantified using dSPM values, yielding noise-normalized statistical maps. Significant alterations contrasted by time were visualized in each patient using a dSPM = 3.3 threshold corresponding to *Z* ≈ 3.3 and *p* < 0.001.

#### EEG sensitivity and craniotomy breach effects

At the post-operative timepoints, the skull defects may cause a ‘breach effect’ that can alter the EEG signal near the craniotomy location^6^. This localized breach effect is known to disrupt mu-alpha EEG frequencies post-craniotomy, dissipating as the bone heals. We studied the sensitivity maps of the EEG signal in both hemispheres at the assessment timepoints. These were observed to be comparable in the ipsilateral and contralateral hemisphere (Suppl. Fig. 2), suggesting minimal impact of the breach effect on the data. We also ensured that reported effects do not depend solely on EEG mu-alpha effects in the ipsilateral hemisphere.

#### Control participant EEG data

Data from twenty healthy control participants who completed the same task with EEG recordings for a separate project were used as a point of reference for the patients (average age: 19.26 years, SD = 1.8 years, 11 female). These data were obtained with a 64-channel Biosemi Active2 EEG system. The participants’ source space was constructed from FreeSurfer (fsaverage) and electrode locations were digitized using the Polhemus Patriot 6DOF motion tracking system and Cortech Locator software. All other parameters were the same as in the patients.

#### Effective connectivity analyses

Time-domain conditional state-space Granger causality (CGC) was computed on source-reconstructed time series using a state-space Granger causality algorithm. CGC was computed using the MVGC toolbox^7^ on per-trial epochs between -115 and 850 ms of the target word presentation onset, first downsampled to 1 kHz and normalized in the range [0,1] to standardize signal amplitude across channels. Pairwise directed interactions between nodes were estimated using the conditional form of Granger causality to account for possible confounding causal associations via intermediate nodes and to mitigate indirect influences from common inputs. CGC was measured using Granger's *F* statistic. Differences in CGC between conditions were defined as the mean across patients of log(*F* post) - log(*F* pre), a formula chosen to stabilize the variance of the long-tailed exponential *F* distribution. To visualize patient-combined CGC changes, data were masked to retain only those edges exhibiting consistent CGC changes (increases or decreases) across all three patients.

#### Intrinsic neurophysiological biomarkers: Time Constant (*τ*_INT_)

For both the intracranial and EEG datasets, we analyzed, respectively, the 1 sec (-1000 to 0 ms relative to sentence onset) and 1.5 sec (-1250 to –250 ms relative to sentence onset) period of silence between the behavioral testing trials. In all three patients, the EEG signal was extracted from the four regions-of-interest (ROIs)—Heschl's Gyrus (HG), Superior Temporal Gyrus (STG), IFG pars opercularis (IFGop), and IFG pars triangularis (IFGtri)—across both hemispheres (ipsilateral and contralateral to the site of resection), across the three timepoints. The intracranial recording data were only available from the ipsilateral hemisphere, and P3 only had electrode coverage of HG and STG (Fig. 1C). The same ROIs were analyzed in the control participant source-localized EEG datasets (*n* = 20). For each trial, the neural signal was mean-centered, and the normalized autocorrelation function (ACF) was computed over lags 0–100 ms. The ACF was then fit with the following exponential decay model:

ACF(τ) = a · exp(−x / τ) + b	(Eqtn. 2)

where *a* is the amplitude, *b* is a baseline offset, *x* is the lag in samples, and *τ* is the time constant (in milliseconds). Curve fitting used SciPy's^8^ curve_fit with parameter bounds for *a* ∈ [0, 2], *b* ∈ [−1, 1], τ ∈ [1, 200] ms, and a maximum of 5000 fits. Fits were accepted only if the coefficient of determination *R*² ≥ 0.50. This threshold excluded fewer than 0.04% of trials, and the mean *R*² across accepted fits was 0.88, indicating high overall model adequacy.

To characterize distributional changes in *τ*_INT_ beyond central tendency, we generated hemisphere-specific quantile-quantile (Q-Q) comparisons of the trial-level tau distributions across timepoints (Fig. 4H). For each hemisphere (ipsilateral and contralateral), empirical *τ*_INT_ quantiles were computed over probability levels from 0.05 to 0.95 for each pairwise comparison (pre-operative vs 2 months, pre-operative vs 6+ months, and 2 months vs 6+ months). In the Q-Q panels (Suppl. Fig. 8), the earlier/reference timepoint was plotted on the y-axis and the later/comparison timepoint on the x-axis, such that deviation from the identity line indicated a distributional shift between timepoints. To quantify uncertainty, we used 1,000 bootstrap resamples within each hemisphere, resampling with replacement while preserving pooled composition across patient and region strata. For each bootstrap sample, we recomputed the quantiles and the quantile difference curve, defined as Q_comparison(*p*) - Q_reference(*p*), and summarized the mean difference with 95% bootstrap confidence intervals across quantile levels. This approach allowed us to visualize whether post-operative changes were concentrated in specific parts of the distribution or reflected broader distribution-wide shifts.

#### Spectral Domain Aperiodic Slope (χ_SPEC_)

Power spectra were estimated from the same single-trial data analysis windows used for the time constant, using Welch's method (1000 sample segments, with a 1000 Hz sample frequency — equivalent to 1 second — with 50% overlap and a Hann window frequency resolution of 0.5 Hz). Spectral parameterization was performed using the FOOOF algorithm (Fitting Oscillations & One Over F; ^9^), applied over the range 3–30 Hz. The aperiodic (1/*f*) component was modelled in *fixed* mode (no spectral knee), and the exponent (representing the spectral slope in log frequency) was the primary parameter of interest. Periodic (oscillatory) components were identified as peaks above the aperiodic fit (peak threshold: 2 SD; peak bandwidth limits: 0.5–12 Hz; maximum 6 peaks). As with the time constant analysis, a minimum goodness-of-fit threshold of *R*² ≥ 0.50 was applied to all FOOOF model fits.

#### Statistical Analyses

For the brain-wide EEG signal testing (Fig. 2), a linear mixed-effects model was fitted to examine the effects of Time period, Hemisphere and Patient on the average regions-of-interest dSPM values between 140 to 165 ms after the target word onset, focusing on the post-initial sound driven response (peaking around ~100 ms). The model included all interaction terms and a random intercept for region of interest (ROI) with three factors, as follows: Time period (3 levels: pre-surgery, 2 months post-surgery, 6+ months post-surgery), Hemisphere (2 levels: ipsilateral, contralateral), and Patient (3 levels: P1, P2, P3). This model was fit with *lme4* using the following *lmer*() notation in *R*:

dSPM value ~	Time period * Hemisphere * Patient + (1 | ROI)	(Eqtn. 3)

For the Region of Interest (ROI) analyses, significance testing was conducted with cluster permutation tests to identify the time periods where the patients overlapped in their significant clusters (*p* < 0.05, cluster length > 5 ms).

Pairwise Conditional Granger Causality (CGC) was computed for every ROI-ROI edge in each patient (see above). At the level of individual patients, changes in effective connectivity were masked by thresholding to include only the top 10% strongest edges based on the between-condition difference in their logged Granger’s *F* statistic, intended to minimize the impact of any spurious results by excluding the weakest changes in CGC. This produced, for each patient, a masked directed graph of connectivity changes (DGCC). Group-level CGC graphs of between-condition differences were masked by intersecting edges of identical sign (all positive/all negative) across all patients’ individual unmasked DGCCs, reflecting consistent directionality of effects across patients.

Statistical inference for both the time constant (*τ*_INT_) and the spectral aperiodic slope (χ_SPEC_) was performed using LME modeling with dependent variables: τ (ms) or aperiodic (1/*f*) exponent, including fixed effects of Hemisphere (ipsilateral vs. contralateral), ROI (HG, STG, IFGop, IFGtri), and Time period (pre surgery, 2 months post, 6+ months post), as follows:

Biomarker ~ Hemisphere * ROI * Time + (1 | Patient)	(Eqtn. 4)

To adjust for the large biomarker sample sizes (per trial, region, hemisphere, and patient), we applied a stringent Bonferroni correction with an acceptance threshold of *p* < 0.001. For tests of differences in the distribution of *τ*_INT_ values, we used the Bowley skewness statistic, a robust quartile-based measure, *B* = (*Q*3 + *Q*1 − 2·*Q*2) / (*Q*3 − *Q*1), where *Q* is the quantile level computed for each session's distribution of *τ*_INT_ values. Differences in Bowley skewness between timepoints were tested using a permutation test (10,000 permutations, two-sided *p*-value). Bootstrap confidence intervals for skewness estimates were derived from 5,000 resamples. Hemisphere-specific (ipsilateral vs. contralateral) skewness was evaluated separately with the same permutation framework. Statistical testing results tables are provided in Suppl. Tables 3-7.

### Supplementary Results

Here we report the complete results associated with the manuscript. There may be some redundancy with the abbreviated version in the manuscript.

Comparison of pre- versus post-surgical T1-weighted MRI anatomical scans were used to identify the surgical lesion location. We observe the involvement of the temporal pole in all three patients, with resection of other brain areas in P1 and P3 (Fig. 1C-D; Methods).

#### Surgical impact on neuropsychological assessment

In comparison to normative data, we evaluated the pre- and 2-month post-operative neuropsychological testing results using the Reliable Change Index (Suppl. Table 2). Post-operatively, P1 and P2 showed significant improvement in short-term memory for faces (WMS-III Faces I; RCI: P1: 3.402, P2: 2.91), whereas P3 showed a significant improvement on the WAIS Digit-Symbol coding task, a measure of processing speed, which could be attributed to improved cognitive function following resection. No other tests showed significantly altered cognitive function in the three patients comparing the pre- versus 2-month post-operative assessments.

#### Surgical impact on semantic expectancy task

Participants completed the semantic expectancy task pre-operatively, 2 months post-operatively, and again at a long-term follow-up assessment (6+ months post-operatively; 6–14 months, testing variability due to COVID-19-related logistical constraints).The behavioral results confirmed stable integration of bottom-up and top-down cues, with all three patients pre-operatively showing strong sensitivity to VOT (P1-3, all *p* < 0.001; complete statistical results in Suppl. Table 3). All three patients also showed significant effects over time (all *p* < 0.001). The two left hemisphere patients’ task performance was significantly degraded pre- versus post-operatively at both 2 and 6+ months (P1 and P2, all *p* < 0.001). The right hemisphere patient’s behavior was unaffected pre- versus 2 months post-operatively but was significantly different in the pre- versus 6+ month assessment period and between the 2 and 6+ months assessment periods (*p* < 0.001; see P3 in Fig. 1E and Suppl. Table 3). All patients performed well on the control tasks in which there was no lexical competition and when the semantic context was never incongruent with the VOT (Suppl. Table 3).

#### Source-localized EEG and effective connectivity impact

We relied on the source-localized EEG results in the patients to identify brain-wide patterns that were reduced or enhanced after ATL resection by comparing the patients’ pre- and post-surgical EEG responses to the target words across the timepoints (Fig. 2A-B). The whole brain results show largely ipsilateral hemisphere enhancement, rather than post-operative reductions, to the target word in all patients (red colormap in Fig. 2A-B; individual patient 3.3 dSPM thresholds, ~*p* < 0.001 uncorrected; Methods). These brain-wide effects were summarized by combining the ipsilateral and contralateral brain regions into a linear mixed-effects model (Fig. 2C). The result shows significant post-operative enhancement in the target word response in both hemispheres in all three patients pre- versus 2 months post-operatively (all *p* < 0.01, excepting the ipsilateral hemisphere in P3 which was not significant; Suppl. Table 4). Figure 2D shows the post-operative alterations in target word responses within the putative compensatory network (e.g., auditory cortex: HG, STG; inferior frontal gyrus: IFGop, IFGtri), revealing similar disruption of low frequency responses to the target word in the ipsilateral hemisphere as reported previously for P1 and P2^1^ and shown in P3 (Suppl. Fig. 3; Suppl. Fig. 4 shows the impact on the semantic expectancy signal in HG). As expected, the patients’ pre-operative EEG signal was not entirely within the normative range defined by controls (Suppl. Fig. 5). The patients’ pre-operative data is therefore an important within-subjects baseline.

We also evaluated effective connectivity during the target word response across the pre- and post-operative EEG data using state-space Conditional Granger Causality analyses. Figure 3A shows the group-mean and individual patient differences in log CGC values, reflecting pre- versus post-operative changes in effective connectivity, with information flow from and to the core set of regions of interest in both hemispheres. We observe a combination of increases and decreases in effective connectivity across time, not specific to either hemisphere. Fig. 3B shows the same group-mean changes in effective connectivity masked by sign to retain only those changes that were directionally consistent across all three patients. At both 2 and 6+ months post- versus pre-surgery, we observe consistent changes in the patients, showing *decreases* in effective connectivity (blue edges; at 2 months, total decreased intra-hemispheric edges: 8 of 16 masked edges), which were more intra-hemispheric at 6+ months (14/18). Comparing 6+ to 2 months post-operatively, we observe marked increases in inter-hemispheric connectivity between these two post-operative periods (red edges). Nearly all consistent increases in effective connectivity during this period are inter-hemispheric in nature (inter-hemispheric CGC increases: 7 of 8 edges).

#### Comparing immediate and longer-term impact on intrinsic neurophysiological biomarkers

The first biomarker analysis focused on calculating the slope of the spectral aperiodic component χ_SPEC_ (Fig. 4A). A steeper aperiodic slope has been associated with a greater contribution of inhibition relative to excitation within the measured neurophysiological signal^10^. Intraoperatively, the spectral slope was steeper in the patients relative to the EEG data in controls (Fig. 4C; all *p* < 0.001 corrected; Suppl. Table 5). This aperiodic slope difference may reflect changes in excitability driven by anesthetic management around the required awake testing periods, with varying effects observed across the three patients (P1 decreased; P2 not significant; P3 increased; see Suppl. Table 5). By comparison, pre-operative EEG aperiodic slope was within the range of the control participants, but following the procedure, there was a gradual increase in the aperiodic slope in the two post-operative assessment periods in all three patients, but only in the ipsilateral hemisphere (Fig. 4D; pre- versus 2 months post-operatively: *p* < 0.001 corrected; Suppl. Table 5; by patient plots: Suppl. Fig. 6). These observations are unlikely to be a craniotomy ‘breach effect’ on the EEG signal^11^, because EEG sensitivity was comparable across hemispheres (Suppl. Fig. 2), and the spectral-slope changes cannot be attributed solely to increases in the mu-alpha frequency range (Fig. 4E).

Next, we evaluated *τ*_INT_, the time-domain intrinsic biomarker associated with neuronal timescales of processing^11^. This analysis relies on calculating signal self-similarity over time, defined as the time constant of the autocorrelation function decay (Fig. 4B). Intraoperatively, the range of observed time constants was within that of controls (Fig. 4F; *p* > 0.05 corrected; Suppl. Table 6). However, as seen with the differential impact on the aperiodic slope (Suppl. Table 5), intraoperatively, the time constant effects differed in each of the patients after surgery (i.e., increased in P1, decreased in P2, no change in P3; Fig. 3F patient-specific results; all *p* < 0.05 corrected; Suppl. Table 6). Perioperatively, the EEG-derived time constant was within the control participant range (Suppl. Table 6), but post-operatively, there was a substantial loss of short time constants, seen as a change in the shape of the distribution and a reduction in the lower quartile in the ipsilateral hemisphere (Fig. 4G). This is seen as positive bias in the Quantile-Quantile plot (Fig. 4H; Bowley distribution skewness test, *p* < 0.05; Suppl. Table 7; Suppl. Fig. 8), where post-operative values exceeded pre-operative values by approximately 4-6 ms at the same quantile level, largely affecting the *τ*_INT_ values in auditory cortex (Suppl. Fig. 7). Finally, analysis of one complete resting-state fMRI dataset, only possible to obtain in P2, showed no significant differences in *τ*_INT_ (Suppl. Fig. 9), unlike the EEG effects for this and all three patients (Fig. 4G; Suppl. Fig. 6).

#### Additional Results: Patient Pre-operative Behavioral Performance in Detail

As we have noted previously^1^, Patient 1 adopted a strategy across both conditions (c.f. Manuscript Fig. 1E and Supplementary Fig. 1) that was more heavily based upon preceding sentential context, with an overall shift towards the /p/ bias. Conversely, Patient 2 retained some ability to integrate voice onset time, but this was diminished from the pre-surgical state, and they showed a new bias in the opposite direction towards /b/, incongruent with the preceding sentential context. This bias cannot be easily explained as a response button bias, given their good performance on the control tasks (see next section). The form of the post-surgical behavioral impact was different between the two patients, potentially because of the more extensive additional left hemisphere medial temporal lobe resection in the case of P1, conducted after the post-disconnection recordings. Patient 3 was largely within the normative range defined by controls until the second assessment period (6+ months). All three patients showed a floor effect in their /b/ bias results. Therefore, they were not fully normative in their performance on the task pre-operatively, as is often the case with epilepsy pathology affecting language and memory performance. The overall behavioral results indicate that the surgical procedure exacerbated an impairment in the ability of both left-hemisphere patients to adaptively integrate VOT information with semantic expectations from sentential context, amidst otherwise intact attentional or general meaning-related abilities, as assessed by the control tasks.

#### Additional Results: Patient Behavioral Performance on Control Trials

Before and after the surgery, the three patients’ performance was nearly perfect on filler trials where the target word was always consistent with the priming sentence and the non-target word was not in lexical competition with it (e.g., “This wall needs another coat of paint” has no /b/ word counterpart; pre-surgery: 100% correct performance in all three patients; 2 months after surgery: 98% for P1; 100% for P2; 100% for P3; 6+ months after surgery: 98.41% for P1; 100% for P2; 100% for P3). Their performance on catch trials where the target word was presented visually as a written word alongside a non-target word was also very high (pre-surgery: 94% for P1; 98% for P2; 96.83% for P3; 2 months after surgery: 92% for P1; 98% for P2; 93.65% for P3; 6+ months after surgery: 92.06% for P1; 97.5% for P2; 96.83% for P3).

### Supplementary Figures

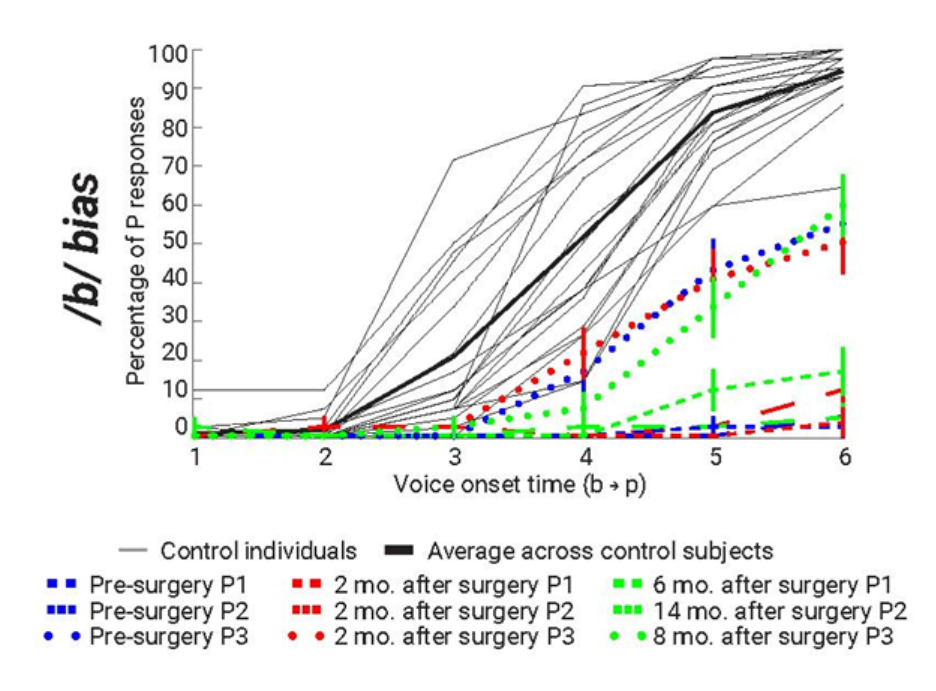

#### Supplementary Figure 1. Additional behavioral results: /b/ bias.

Percentage of P behavioral responses to the /b/ bias words in relation to the VOTs (from /b/ to /p/ sounds) from the control participants and Patients 1, 2, and 3, approximately one month before, two months, and 6+ months after the surgery. Format as in manuscript Fig. 1E. The floor effect pre- and post-disconnection on the /b/ bias condition for the patients precludes drawing conclusions about the impact or lack thereof on this condition (see /p/ bias effects in manuscript Fig. 1E).

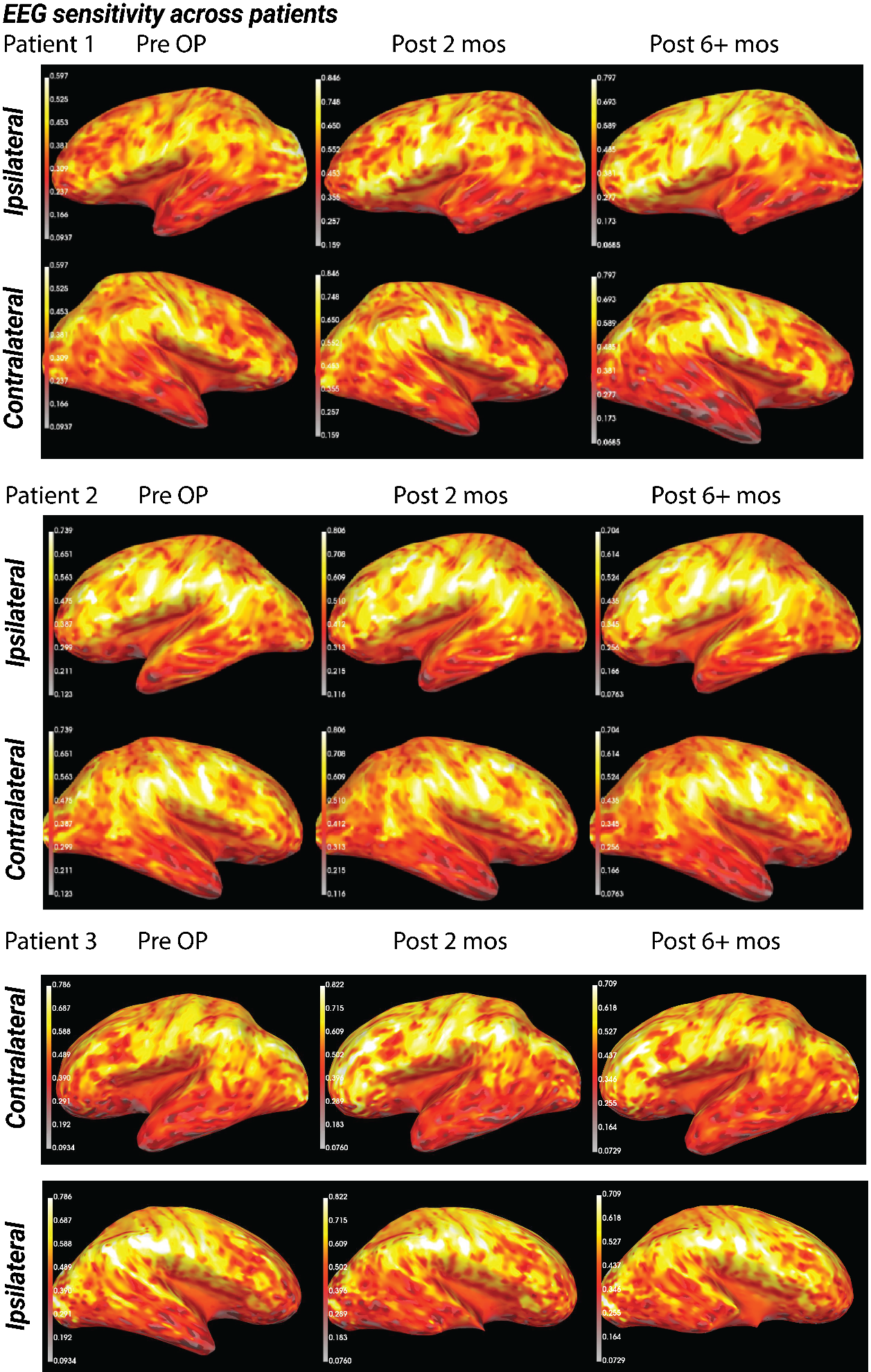

#### Supplementary Figure 2. EEG sensitivity maps across patients.

Sensitivity maps are created from MNE toolbox forward operators that indicate how well different sensor types will be able to detect neural currents from different regions of the brain. Each patient’s sensitivity maps are plotted separately for both hemispheres. Yellow colors show the most sensitive areas, and red shows the least. If the breach effect was apparent for the patients, the sensitivity maps should show a larger sensitivity over the craniotomies.

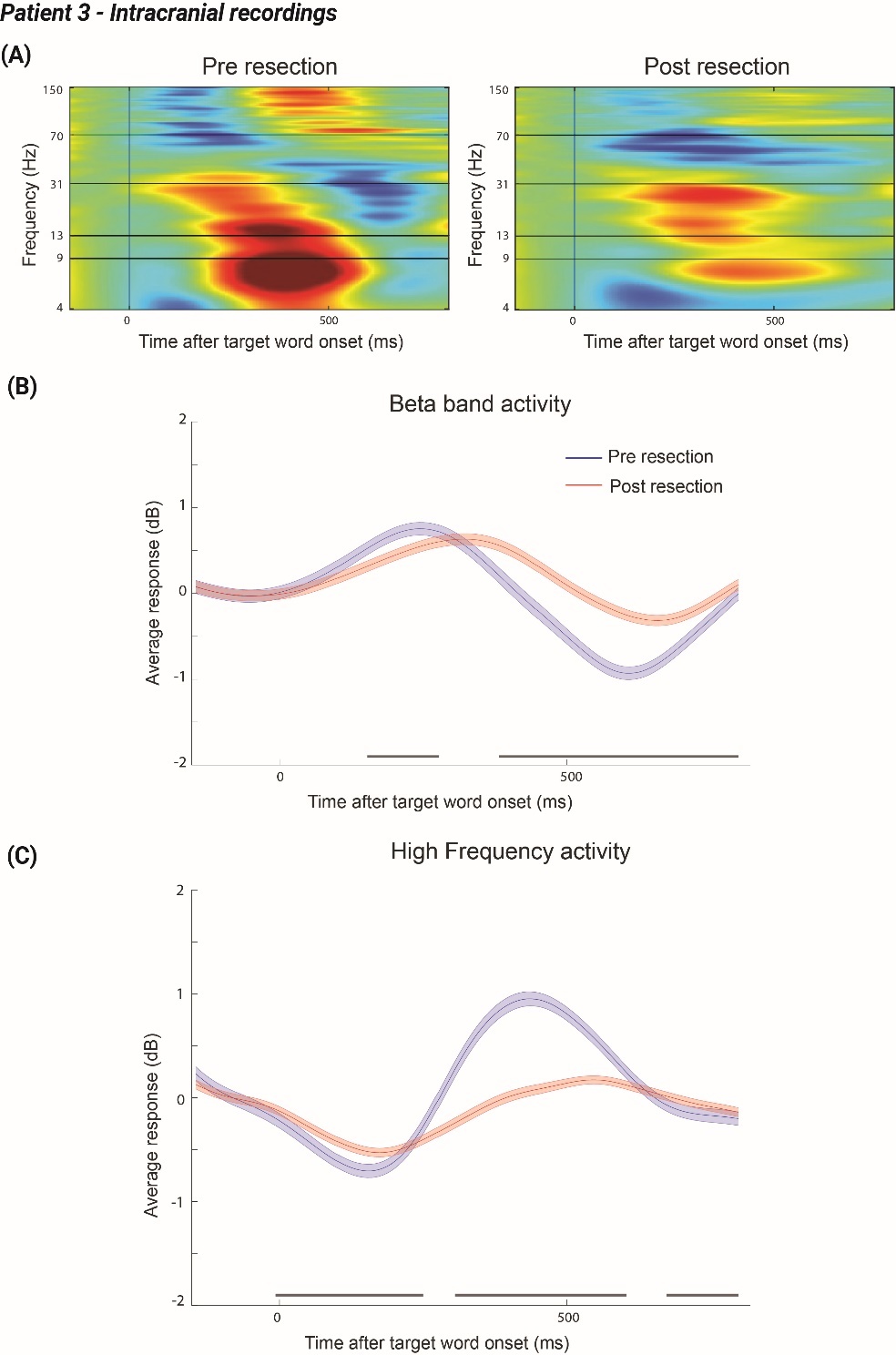

#### Supplementary Figure 3. Intracranial recording responses to the target word in Patient 3: Heschl’s gyrus.

(A) Pre- and post-resection intracranial recording responses to the target word in Patient 3, in the format of P1 and P2 reported previously^1^. Similarly to P1 and P2, we observe lower frequencies (<30 Hz, such as beta) showing disruption in the target word response post-resection (B). However, the high-frequency activity magnification reported in P1 and P2 is absent in this patient (namely, high gamma, 70-150 Hz). Average responses are plotted together with the standard error of the mean. Gray bars below the plots in (B-C) are cluster significant effects (see Methods).

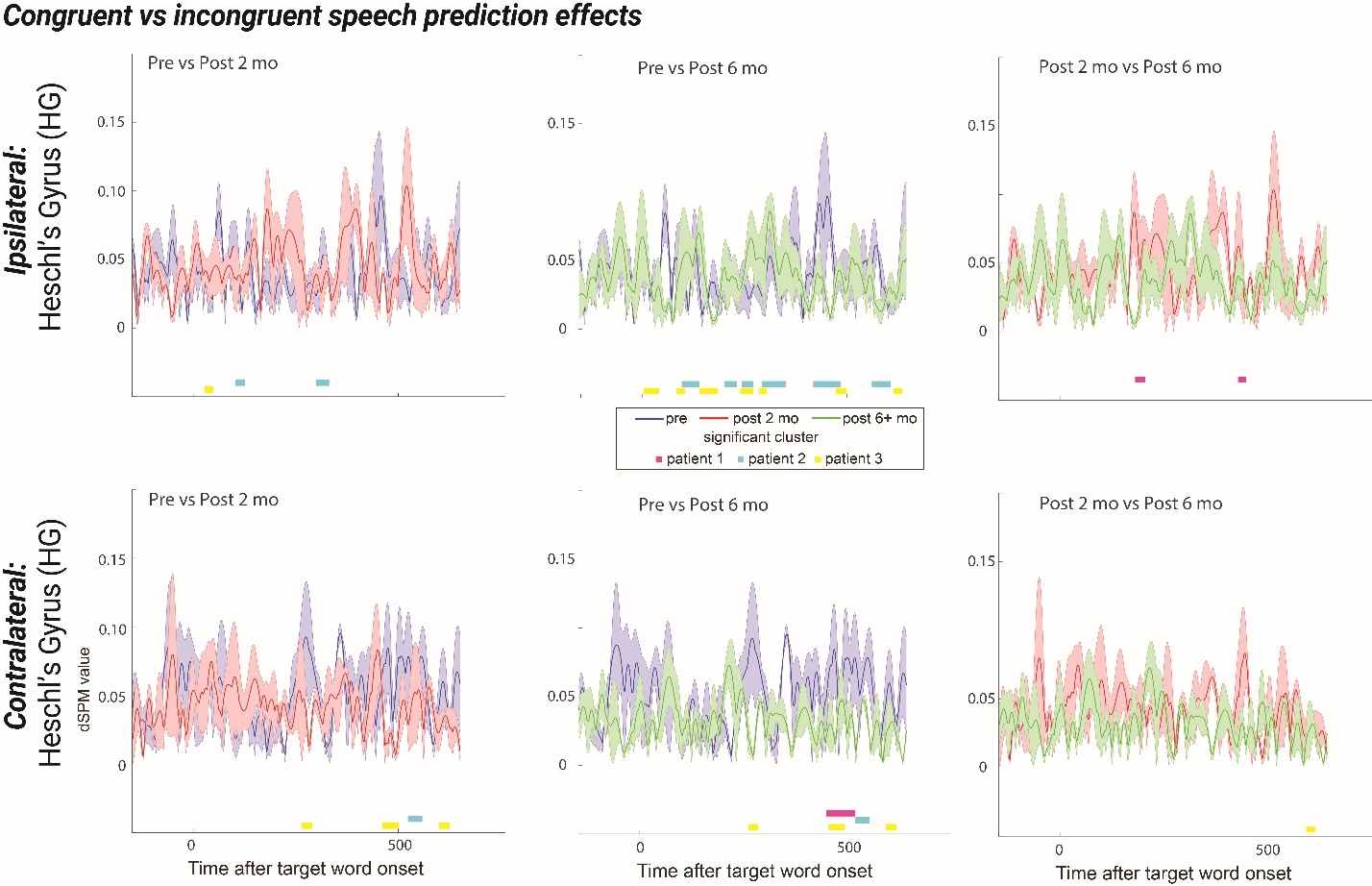

#### Supplementary Figure 4. Congruent versus incongruent target word EEG responses pre- and post-surgery in Heschl’s gyrus.

Activation is compared between patients’ responses to the congruent and incongruent conditions, averaged over the 3 patients. The difference wave is calculated as Congruent – Incongruent responses. Pre-surgical responses are shown in blue, 2 months post-surgical responses are in red, and 6+ months post-surgical responses are in green, plotted together with the standard error of the mean (SEM) between trials. Cluster-based permutation testing was conducted within each patient. Significant (*p* < 0.05) differences between the responses recorded at the two time periods are pictured underneath the line plots. The clusters are signaled by magenta for P1, cyan for P2, and yellow for P3.

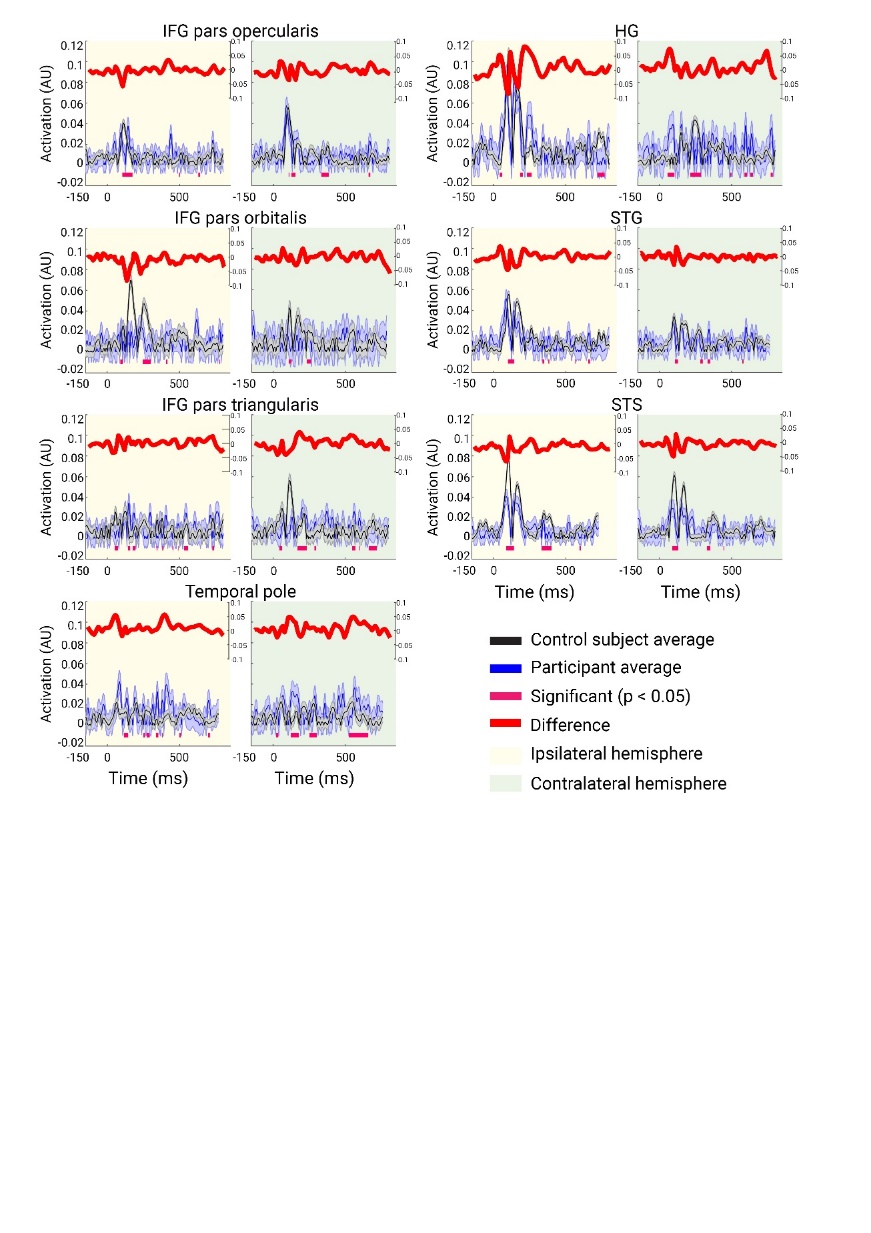

#### Supplementary Figure 5. Average source-localized EEG activation of regions of interest comparing patients and control participants (*n* = 20).

Activation is compared between patients’ pre-surgical activations (blue line plots) vs. control participants’ activation (black line plots), plotted together with the standard error of the mean (SEM) between trials and with the difference (3 patients’ average minus control participants’ average) between the two (red line). The left columns (yellow background) denote the ipsilateral, and the right columns (green background) denote the contralateral activation. The lines under the plots show statistically significant (*p* < 0.05) differences between the averaged control participant and the averaged patient responses, based on cluster-based permutation testing.

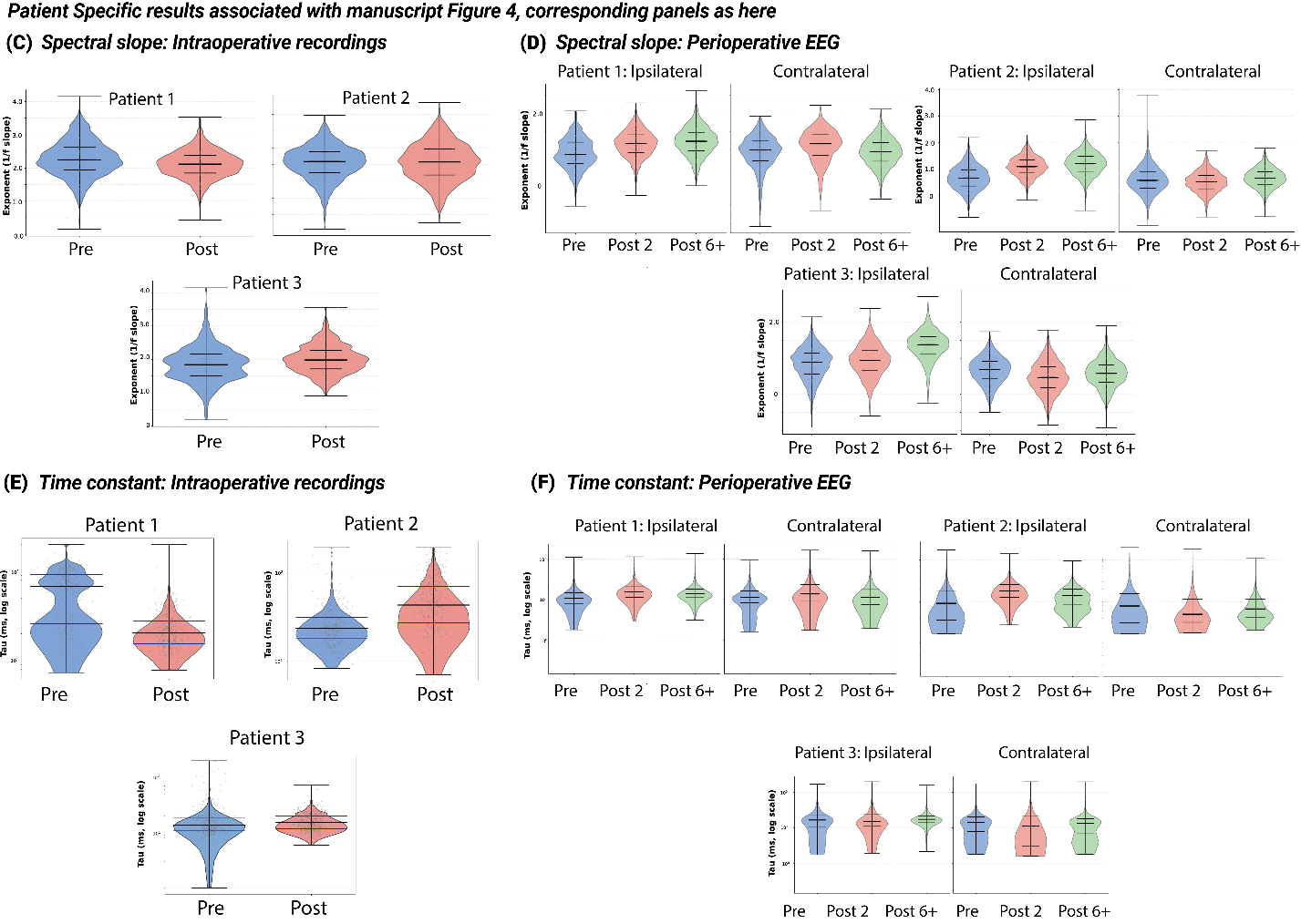

#### Supplementary Figure 6: Aperiodic slope and time constant results – by patient.

This is the patient-specific complement of manuscript Figure 4, with panel labels consistent with the manuscript figure. As a result, this figure starts with panel C, given that manuscript Figure 4A-B were example biomarker analysis plots not needed here. (C) This complements the combined data in Figure 4C which shows pre- and post-operative consistency in the patient-specific spectral slope. (D) This complements the combined data in Figure 4D, which shows that the combined effect of an increased aperiodic slope post-operatively in the ipsilateral hemisphere is also evident in the patient-specific results. (E) These panels are also shown in Figure 4E because the patients differ in the pattern of pre- and post-operative results. (F) This complements the combined data in Figure 4F, showing that the combined effect of skewness (i.e., loss of shorter time constants) in the ipsilateral hemisphere post-operatively is evident in all three patients.

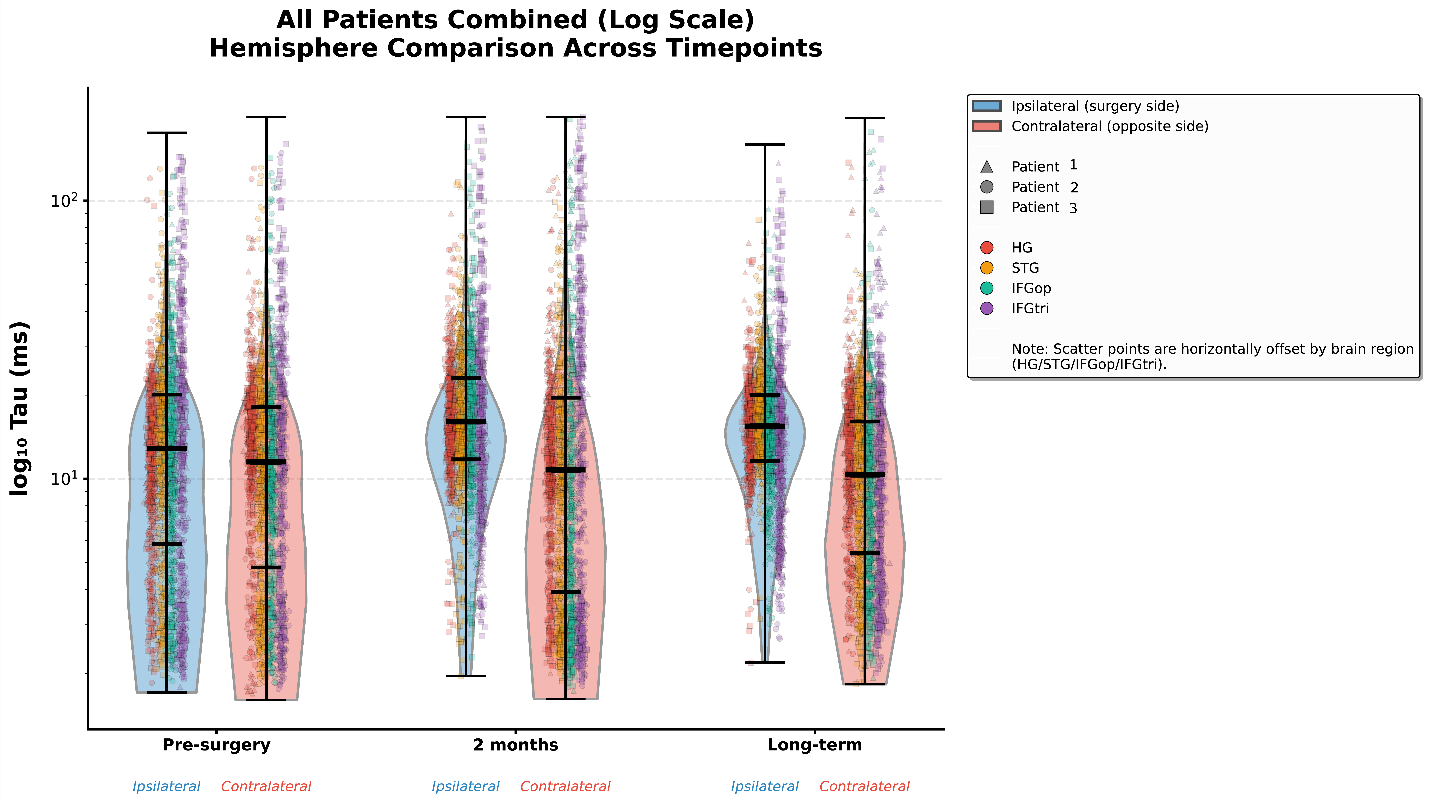

#### Supplementary Figure 7: EEG time constant effects by brain area.

This figure complements Figure 4F, showing the data subdivided by time constants by brain area (HG, STG, IFGop, IFGtri). We note that although there is a shift away from lower time constants post-operatively in the ipsilateral hemisphere, this does not seem to be the case for the IFG, where the distribution of time constants continues to span lower values.

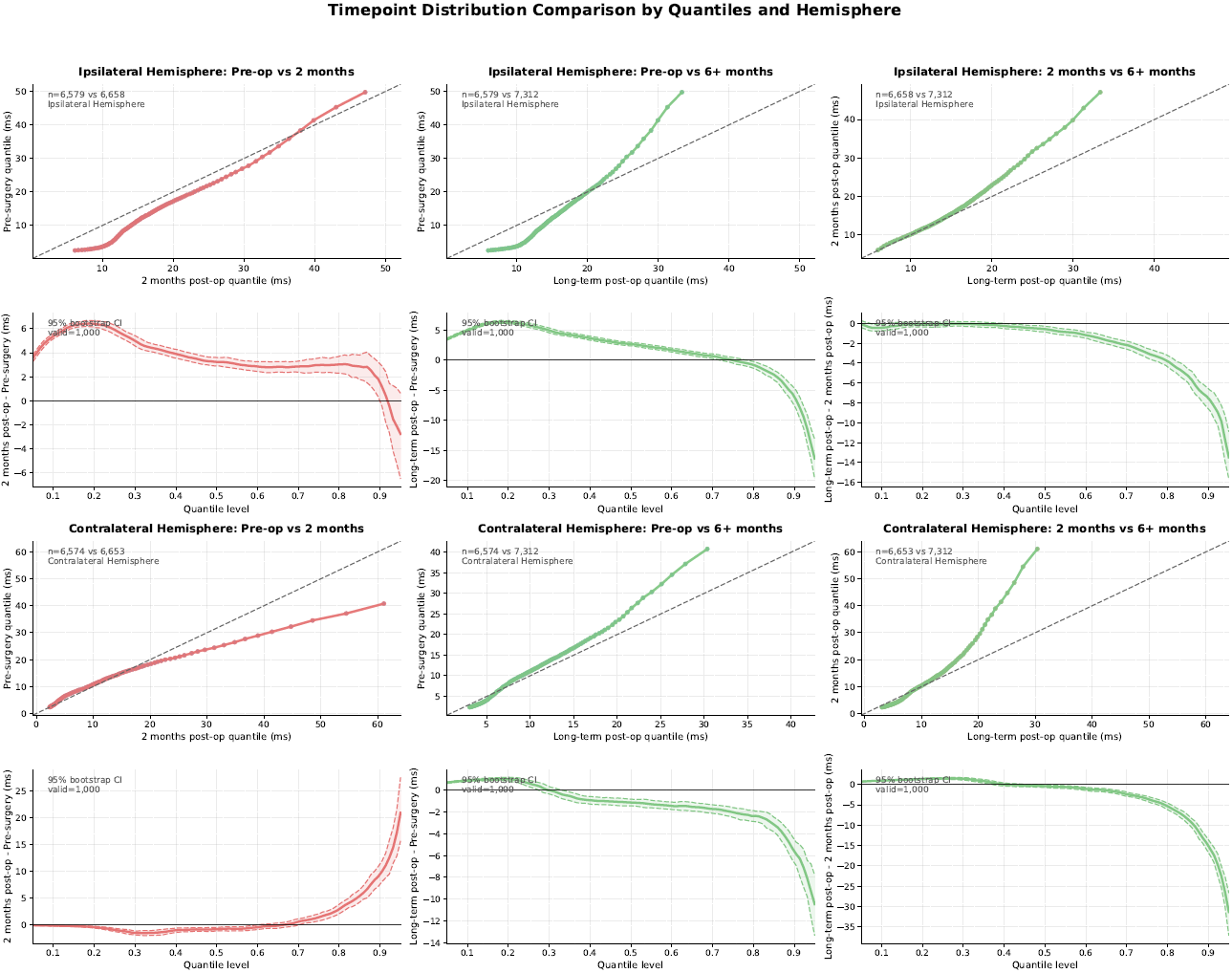

#### Supplementary Figure 8: Time constant distribution skewness Q-Q plots.

This figure complements the manuscript Figure 4 Q-Q plots, here showing the complete set of quartile skewness plots comparing pre versus post-operative skewness via quartile-quartile (QQ) plots. See manuscript text for details.

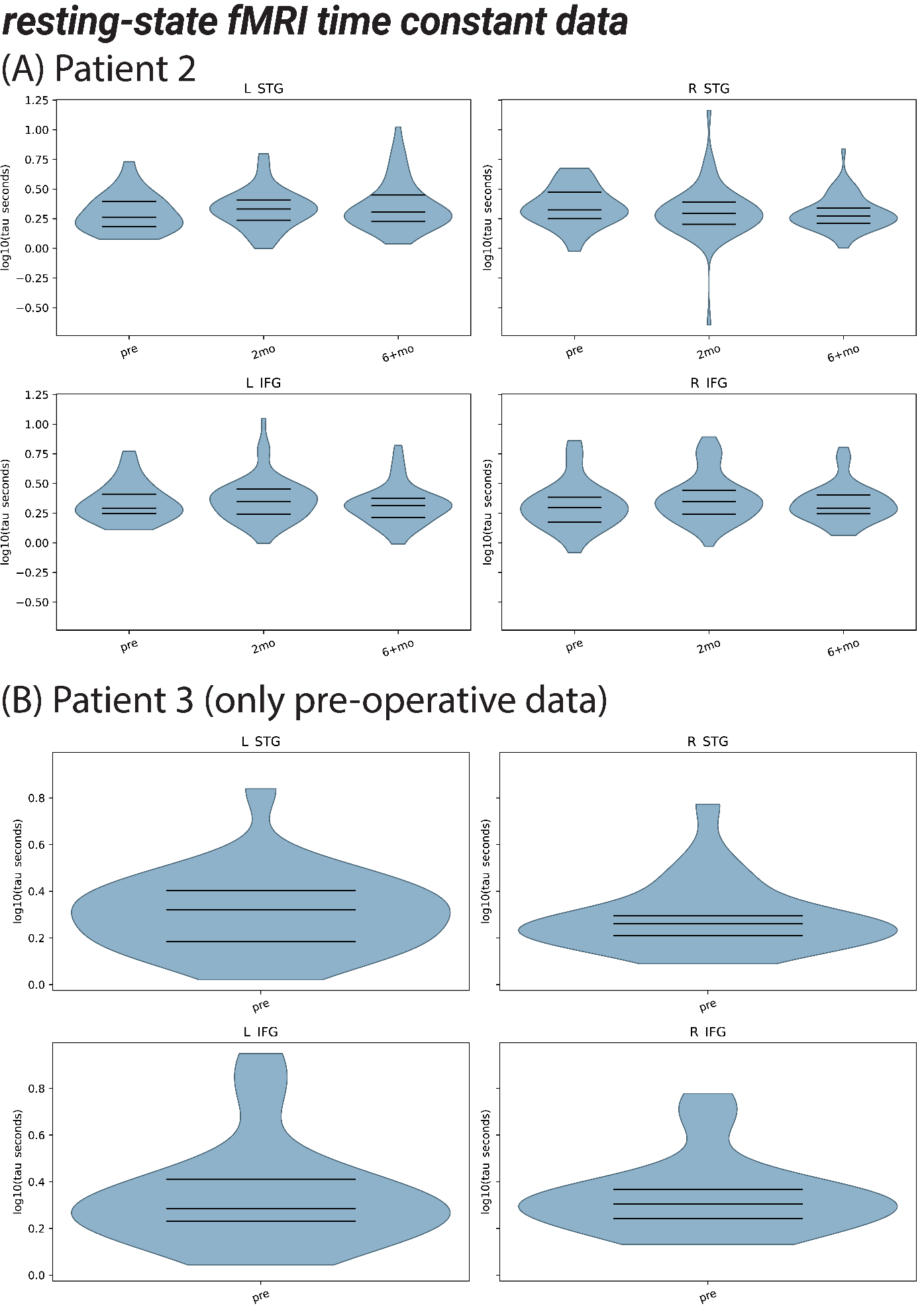

#### Supplementary Figure 9: Resting-state fMRI time constant results.

These were the only resting-state fMRI data obtained, a complete dataset in P2 (A) and only pre-operative rs-fMRI in P3 (B). The rs-fMRI analyses followed the approach previously established for extracting time constants from rs-fMRI data^12^, whereby the rs-fMRI time-varying signal for voxels within the given brain areas (STG and IFG) was processed using the auto-correlation function fit, and time constant extracted as a function of volume time in seconds (TR). Although the time constant variability is expectedly with a median value of around one TR (2.26 s; 7.95 minutes per scanning run), unlike the EEG time constant effects for Patient 2 reported in manuscript Fig. 4G, we do not see post-operative differences in this patient, either in the skewness of the distribution or a shift in the median value of rs-fMRI time constants. Since these results are only a case study, they are appropriate for reporting in the Supplementary Materials.

### Supplementary Tables

#### Supplementary Table 1. The number of trials completed, and the electrodes excluded from analysis for each patient.

Exclusion was based on very high noise levels, which the source-localization method is highly susceptible to.

| **Patient** | **Recording time** | **Number of electrodes discarded from analysis** | **Number of all recorded trials** | **Number of trials after artifact rejection** |
| --- | --- | --- | --- | --- |
| **P1** | Pre-surgery | 4 | 253 | 246 |
|  | Post-surgery – 2 months | 3 | 504 | 504 |
|  | Post-surgery – 6 months | 19 | 504 | 481 |
| **P2** | Pre-surgery | 7 | 504 | 472 |
|  | Post-surgery – 2 months | 3 | 360 | 354 |
|  | Post-surgery – 14 months | 13 | 504 | 504 |
| **P3** | Pre-surgery | 0 | 504 | 496 |
|  | Post-surgery – 2 months | 1 | 504 | 465 |
|  | Post-surgery – 8 months | 0 | 504 | 504 |

#### Supplementary Table 2. Pre- and post-operative neuropsychological testing scores and calculated Reliable Change Index (RCI).

Test abbreviations are as follows: Rey Auditory Verbal Learning Test (RAVLT), Wechsler Adult Intelligence Scale 4th Edition (WAIS-IV), Wechsler Memory Scale 3rd Edition (WMS-III), Controlled Oral Word Association Test (COWA), Trail Making Test (TMT), Boston Naming Test (BNT), Beck Depression Inventory - II (BDI-II), and Beck Anxiety Inventory (BAI). Significant RCI values are indicated in bold italic type.

|  | **Test** | **Subtest** | **Pre-operative score** | **Pre-operative**  **Z-score** | **Post-operative score** | **RCI** |
| --- | --- | --- | --- | --- | --- | --- |
| **P1** | RAVLT | Trial 1 | 5 | -0.563 | 4 | -0.533 |
|  |  | Recall | 8 | -0.200 | 5 | -1.038 |
|  | WMS-III | Logical Memory I | 27 | -1.000 | 25 | -0.921 |
|  |  | Logical Memory II | 12 | -1.000 | 14 | 0.788 |
|  |  | Faces I | 31 | -1.000 | 38 | ***3.402*** |
|  |  | Faces II | 29 | -1.000 | 33 | ***1.994*** |
|  |  | Spatial Span | 12 | -1.000 | 16 | 1.332 |
|  | TMT | A | 36 | 0.323 | 25.4 | -0.966 |
|  |  | B | 56 | -1.194 | 69.8 | 0.440 |
|  | BDI |  | 18 | N/A | 23 | N/A |
| **P2** | WAIS-IV | Similarities | 9 | -0.333 | 12 | 1.348 |
|  |  | Digit Span | 9 | -0.333 | 9 | 0.000 |
|  |  | Arithmetic | 11 | 0.333 | 13 | 1.090 |
|  |  | Digit Symbol Coding | 8 | -0.667 | 10 | 1.547 |
|  | RAVLT | Trial I | 7 | 0.167 | 7 | 0.000 |
|  |  | Recall | 10 | -0.429 | 8 | -0.692 |
|  | WMS-III | Logical memory I | 41 | 0.000 | 39 | -0.921 |
|  |  | Logical Memory II | 23 | 0.000 | 15 | ***-3.155*** |
|  |  | Faces I | 25 | -1.667 | 31 | ***2.916*** |
|  |  | Faces II | 30 | -1.333 | 31 | 0.498 |
|  | COWA |  | 41 | 0.418 | 56 | 1.907 |
|  | BNT |  | 55 | -0.580 | 57 | 0.722 |
|  | TMT | A | 21 | -0.390 | 30 | 0.820 |
|  |  | B | 43 | -0.621 | 61 | 0.472 |
|  | BDI |  | 30 | N/A | 8 | N/A |
|  | BAI |  | 30 | N/A | 17 | N/A |
| **P3** | WAIS-IV | Processing Speed Index Standard Score | 94 | -0.4 | 111 | N/A |
|  |  | Block design | 13 | 1 | 16 | 1.333 |
|  |  | Digit Span | 12 | 0.67 | 15 | -0.527 |
|  |  | Digit Symbol Coding | 8 | -0.67 | 12 | ***2.108*** |
|  |  | Symbol Search | 10 | 0.00 | 12 |  |
|  | RAVLT | Trial 1 | 6 | -0.39 | 4 | 0.00 |
|  |  | Recall | 9 | -0.79 | 9 | 0.00 |
|  | COWA |  | 50 | 1.01 | 54 | 1.653 |
|  | BNT |  | 55 | -0.58 | 55 | 0.00 |
|  | TMT | A | 22 | -0.65 | 18 | -0.064 |
|  |  | B | 45 | -0.82 | 33 | -0.058 |
|  | BDI |  | 6 | N/A | 4 | N/A |

#### Supplementary Table 3. Behavioral statistical test results (associated with Fig. 1E)

This table summarizes the patient-level linear mixed models predicting phoneme categorization (binary /p/ vs /b/ responses) as a function of Voice onset time (VOT), contextual Bias (B or P), and Time period (pre-, 2 months post-, and 6+ months post-surgery). Estimates are shown on the log-odds scale. Interaction effects are reported as likelihood ratio tests unless otherwise noted. Result labels are based on corrected p-values: *** p < 0.001, ** p < 0.01, * p < 0.05, n.s. non-significant.

**Main effects and interactions from GLMM analyses (binomial logistic models)**

| **Effect** | **P1** | **P2** | **P3** |
| --- | --- | --- | --- |
| **VOT** | *β* = 1.02, *SE* ≈ 0.00, *z* = 541.2, *p* < 0.001 *** | *β* = 1.22, *SE* = 0.19, *z* = 6.60, *p* < 0.001 *** | *β* = 3.78, *SE* = 0.37, *z* = 10.15, *p* < 0.001 *** |
| **Bias** | *β* = 6.09, *SE* ≈ 0.00*, z* = 3232, *p* < 0.001 *** | *β* = 3.51, *SE* = 0.33, *z* = 10.68, *p* < 0.001 *** | *β* = 3.29, *SE* = 0.25, *z* = 13.32, *p* < 0.001 *** |
| **Time** | *χ²(2)* = 763.74, *p* < 0.001*** | *χ²(2) =* 53.75, *p* < 0.001*** | *χ²(2)* = 16.03, *p* < 0.001*** |

**Time contrasts (log-odds scale)**

| **Contrast** | **P1** | **P2** | **P3** |
| --- | --- | --- | --- |
| **Pre vs 2 months post** | *β* = 0.198, *SE* ≈ 0.00, *z* = -858.98, *p* < 0.001 *** | *β =* 6.18, *SE* = 1.753, *z* = 6.42, *p* < 0.001 *** | *β* = 0.13, *SE* = 0.262*, z* = 0.574, *p* = 1 n.s. |
| **Pre vs 6+ months post** | *β* = 0.7305, *SE* ≈ 0.00, z = -161.293, *p* < 0.001 *** | *β* = 3.745, *SE* = 0.87, *z* = 5.68, *p* < 0.001 *** | *β* = 0.88, *SE* = 0.568, *z* = 3.732, *p* < 0.001 *** |
| **2 months vs 6+ months post** | *β* = 3.686, *SE* ≈ 0.00, *z* = 481.6*, p* < 0.001 *** | *β* = 0.606, *SE* = 0.177, *z* = -1.715, *p =* 0.259 n.s. | *β =* 0.75, *SE* = 0.495, *z* = 3.187, *p =* 0.004 ** |

**Interaction effects (model comparison χ² tests)**

| **Interaction** | **P1** | **P2** | **P3** |
| --- | --- | --- | --- |
| **Bias x VOT** | *χ²(1)* = 0.76, *p* = 0.38 n.s. | *χ²(1)* = 4.61, *p* = 0.032 * | *χ²(1)* = 5.82, *p* = 0.016 * |
| **VOT x Time** | *χ²(2)* = 12.38, *p* = 0.002 ** | *χ²(2)* = 12.35, *p* = 0.002 ** | *χ²(2)* = 4.89, *p* = 0.087 n.s. |
| **Bias x Time** | *χ²(2)* = 1.88, *p* = 0.39 n.s. | *χ²(2)* = 32.2, *p* < 0.001 *** | *χ²(2)* = 8.82, *p* = 0.012 * |

#### Supplementary Table 4. Brain-wide longitudinal EEG statistical testing results (associated with Fig. 2C)

Summary of patient-level linear mixed models predicting the average dSPM activation of regions of interest as a function of Time period (pre-, 2 months post-, and 6+ months post-surgery, Hemisphere (Ipsilateral or contralateral to the surgery). There was a significant main effect of Time period (*χ²(12)* = 108.81, *p* < 0.001). Pre-surgery and 2 months post-surgery were associated with higher values (*β* = 0.010, *SE* = 0.002, *p* < 0.001). A significant main effect of Hemisphere was observed (*β* = 0.006, *SE* = 0.002, *p* = 0.003). There was a significant interaction between Time period and Patient, indicating that the effect of time differed across patients, as shown in the table below and discussed in the manuscript. No significant interactions were found between Time period and Hemisphere, nor was the three-way interaction among Time period, Hemisphere, and Patient significant (all *p* > 0.10). Result labels are based on corrected p-values: *** p < 0.001, ** p < 0.01, * p < 0.05, n.s. non-significant.

|  | **P1** | | **P2** | | **P3** | |
| --- | --- | --- | --- | --- | --- | --- |
| **Time period** | **Ipsilateral** | **Contralat.** | **Ipsilateral** | **Contralat.** | **Ipsilateral** | **Contralat.** |
| **Pre vs 2 months post** | *β* = -0.0096, *p* = 0.0093 ** | *β* = -0.0107, *p* = 0.003 ** | *β* = -0.0188, *p* < 0.001 *** | *β* = -0.0105, *p* = 0.004 ** | *β* = -0.0032, *p* = 0.58 n.s. | *β* = -0.0095, *p* = 0.011 * |
| **Pre vs 6+ months post** | *β* = 0.0059,  *p* = 0.16 n.s. | *β* = 0.0065,  *p* = 0.12 n.s. | *β* = -0.0046, *p* = 0.33 n.s. | *β* = -0.0008, *p* = 0.96 n.s. | *β* = -0.004,  *p* = 0.37 n.s. | *β* = -0.0084, *p* = 0.028 * |
| **2 months vs 6+ months post** | *β* = 0.0155,  *p* < 0.001 *** | *β* = 0.0172,  *p* < 0.001 *** | *β* = 0.0142,  *p* < 0.001 *** | *β* = 0.0096,  *p* = 0.009 ** | *β* = -0.0011, *p* = 0.94 n.s. | *β* = 0.0011,  *p* = 0.94 n.s. |

#### Supplementary Table 5. Aperiodic slope statistical testing results

Intraoperative aperiodic slope comparisons are shown for each patient or patients combined versus the control participant data for spectral slope distribution. Corrected *p*-values are Bonferroni-adjusted across 20 planned tests. Estimate/beta is the mixed-effects coefficient for the comparisons (see manuscript Methods for statistical model). Result labels are based on corrected p-values: *** p < 0.001, ** p < 0.01, * p < 0.05, n.s. non-significant.

**(A) Intraoperative - Comparing control participants’ EEG aperiodic slope central tendency to patients intraoperative aperiodic slope (manuscript Fig. 4C)**

| **Comparison** | **Control mean** | **Patient mean** | **β** | **SE** | **z** | **p** | **Corrected p** | **Result** |
| --- | --- | --- | --- | --- | --- | --- | --- | --- |
| Controls vs patients pre-op | 0.8746 | 2.1141 | 1.1840 | 0.0996 | 11.8879 | 1.37e-32 | 1.37e-28 | *** |
| Controls vs patients post-op | 0.8746 | 2.0723 | 1.1501 | 0.0996 | 11.5507 | 7.32e-31 | 7.32e-27 | *** |

| **Patient** | **Comparison** | **Control mean** | **Patient mean** | **Estimate / beta** | **SE** | **z** | **p** | **Corrected p** | **Result** |
| --- | --- | --- | --- | --- | --- | --- | --- | --- | --- |
| P1 | Controls vs patient pre-op | 0.8746 | 2.2751 | 1.3946 | 0.1656 | 8.4229 | 3.67e-17 | 3.67e-13 | *** |
| P1 | Controls vs patient post-op | 0.8746 | 2.1122 | 1.2317 | 0.1656 | 7.4389 | 1.02e-13 | 1.02e-9 | *** |
| P2 | Controls vs patient pre-op | 0.8746 | 2.0556 | 1.1751 | 0.1662 | 7.0700 | 1.55e-12 | 1.55e-8 | *** |
| P2 | Controls vs patient post-op | 0.8746 | 2.0685 | 1.1879 | 0.1662 | 7.1474 | 8.84e-13 | 8.84e-9 | *** |
| P3 | Controls vs patient pre-op | 0.8746 | 1.8407 | 0.8925 | 0.1660 | 5.3770 | 7.57e-8 | 7.57e-4 | *** |
| P3 | Controls vs patient post-op | 0.8746 | 1.9996 | 1.0513 | 0.1657 | 6.3433 | 2.25e-10 | 2.25e-6 | *** |

**(B) Intraoperative - Pre versus post operative aperiodic slope in the patients (manuscript Fig. 4C)**

| **Patient** | **Comparison** | **Pre mean** | **Post mean** | **β** | **SE** | **z/t** | **p** | **Corrected p** | **Direction / result** |
| --- | --- | --- | --- | --- | --- | --- | --- | --- | --- |
| P1 | Post vs pre | 2.2751 | 2.1122 | -0.1629 | 0.0176 | -9.2393 | 4.69e-20 | 4.69e-16 | decrease *** |
| P2 | Post vs pre | 2.0556 | 2.0685 | 0.0129 | 0.0174 | 0.7410 | 0.459 | 1 | n.s. |
| P3 | Post vs pre | 1.8407 | 1.9996 | 0.1589 | 0.0267 | 5.9550 | 3.37e-09 | 3.37e-05 | increase *** |

**(C) EEG data - Comparing control EEG spectral slope distribution to patients’ EEG spectral slope pre-operatively (manuscript Fig. 4D)**

| **Comparison** | **Control mean** | **Patient pre-op mean** | **β** | **SE** | **z** | **p** | **Corrected p** | **Result** |
| --- | --- | --- | --- | --- | --- | --- | --- | --- |
| Controls vs patients pre-op | 0.8746 | 0.7397 | -0.0995 | 0.1000 | -0.9957 | 0.319 | 1 | n.s. |

| **Patient** | **Comparison** | **Control mean** | **Patient mean** | **Estimate / beta** | **SE** | **z** | **p** | **Corrected p** | **Result** |
| --- | --- | --- | --- | --- | --- | --- | --- | --- | --- |
| P1 | Controls vs patient pre-op | 0.8746 | 0.9315 | 0.0510 | 0.1649 | 0.3091 | 0.757 | 1 | n.s. |
| P2 | Controls vs patient pre-op | 0.8746 | 0.6441 | -0.2364 | 0.1655 | -1.4288 | 0.153 | 1 | n.s. |
| P3 | Controls vs patient pre-op | 0.8746 | 0.7537 | -0.1269 | 0.1650 | -0.7688 | 0.442 | 1 | n.s. |

**(D) EEG data - EEG slope effects in the ipsilateral versus contralateral hemisphere in the patients (manuscript Fig. 4D)**

| **Hemisphere** | **Test / comparison** | **Estimate / statistic** | **SE** | **z / chi-square** | **p** | **Corrected p** | **Result** |
| --- | --- | --- | --- | --- | --- | --- | --- |
| Ipsilateral | Time period omnibus | N/A | N/A | 4269.06 | <1e-300 | 1e-296 | *** |
| Ipsilateral | Pre vs 2 months | 0.2648 | 0.0073 | 36.1467 | 4.19e-286 | 4.19e-282 | *** |
| Ipsilateral | Pre vs 6+ months | 0.4631 | 0.0071 | 65.2996 | <1e-300 | 1e-296 | *** |
| Ipsilateral | 2 months vs 6+ months | 0.1983 | 0.0070 | 28.1971 | 6.35e-175 | 6.35e-171 | *** |
| Contralateral | Time period omnibus | N/A | N/A | 0.667 | 0.717 | 1 | n.s. |

* Note: Below patient specific comparisons reported only for the above main significant omnibus effects (i.e., ipsilateral hemisphere).

| **Patient** | **Hemisphere** | **Comparison** | **Estimate / beta** | **SE** | **z** | **p** | **Corrected p** | **Result** |
| --- | --- | --- | --- | --- | --- | --- | --- | --- |
| P1 | Ipsilateral | Pre vs 2 months | 0.2628 | 0.0134 | 19.6563 | 5.10e-86 | 1.02e-84 | increase *** |
| P1 | Ipsilateral | Pre vs 6+ months | 0.3254 | 0.0136 | 23.9392 | 1.20e-126 | 2.39e-125 | increase *** |
| P1 | Ipsilateral | 2 months vs 6+ months | 0.0627 | 0.0109 | 5.7631 | 8.26e-9 | 1.65e-7 | increase *** |
| P2 | Ipsilateral | Pre vs 2 months | 0.4247 | 0.0126 | 33.8403 | 5.03e-251 | 1.01e-249 | increase *** |
| P2 | Ipsilateral | Pre vs 6+ months | 0.5229 | 0.0114 | 45.6854 | <1e-300 | <2e-299 | increase *** |
| P2 | Ipsilateral | 2 months vs 6+ months | 0.0982 | 0.0127 | 7.7102 | 1.26e-14 | 2.51e-13 | increase *** |
| P3 | Ipsilateral | Pre vs 2 months | 0.0913 | 0.0120 | 7.6047 | 2.86e-14 | 5.71e-13 | increase *** |
| P3 | Ipsilateral | Pre vs 6+ months | 0.5025 | 0.0118 | 42.7332 | <1e-300 | <2e-299 | increase *** |
| P3 | Ipsilateral | 2 months vs 6+ months | 0.4112 | 0.0122 | 33.6147 | 1.02e-247 | 2.04e-246 | increase *** |

#### Supplementary Table 6. Time constant central tendency statistical testing results

Intrinsic time constant central tendency comparisons are shown for each patient or patients combined versus the control participant data for spectral slope distribution. Corrected *p*-values are Bonferroni-adjusted across 20 planned tests. Estimate/beta is the mixed-effects coefficient for the comparisons (see manuscript Methods for statistical model). Result labels are based on corrected p-values: *** p < 0.001, ** p < 0.01, * p < 0.05, n.s. non-significant.

**(A) Intraoperative recording - pre versus post operative time constant central tendency in each patient (manuscript Fig. 4F)**

| **Patient** | **Comparison** | **Pre mean** | **Post mean** | **β** | **SE** | **t** | **p** | **Corrected p** | **Direction / result** |
| --- | --- | --- | --- | --- | --- | --- | --- | --- | --- |
| P1 | Post vs pre | 45.068 | 19.075 | -26.099 | 1.027 | -25.411 | 1.56e-128 | 1.56e-124 | decrease *** |
| P2 | Post vs pre | 28.844 | 45.399 | 16.544 | 1.079 | 15.337 | 4.31e-51 | 4.31e-47 | increase *** |
| P3 | Post vs pre | 19.807 | 16.680 | -3.158 | 0.948 | -3.333 | 8.85e-04 | 1 | n.s. |

**(B) EEG - control compared to patients EEG pre-operative time constant (manuscript Fig. 4G)**

| **Comparison** | **Control mean** | **Patient pre-op mean** | **β** | **SE** | **z** | **p** | **Corrected p** | **Result** |
| --- | --- | --- | --- | --- | --- | --- | --- | --- |
| Controls vs patient pre-op | 23.345 | 16.309 | -7.105 | 4.535 | -1.567 | 0.117 | 1 | n.s. |

| **Patient** | **Comparison** | **Control mean** | **Patient pre-op mean** | **Estimate / beta** | **SE** | **z** | **p** | **Corrected p** | **Result** |
| --- | --- | --- | --- | --- | --- | --- | --- | --- | --- |
| P1 | Controls vs patient pre-op | 23.3454 | 13.3457 | -10.0109 | 7.8043 | -1.2827 | 0.2 | 1 | n.s. |
| P2 | Controls vs patient pre-op | 23.3454 | 14.5877 | -8.7654 | 7.7957 | -1.1244 | 0.261 | 1 | n.s. |
| P3 | Controls vs patient pre-op | 23.3454 | 19.2994 | -4.0547 | 7.8526 | -0.5163 | 0.606 | 1 | n.s. |

**(C) EEG - time constant effects in the patients pre versus post-operatively (manuscript Fig. 4G)**

| **Model / test** | **Effect / comparison** | **Estimate / statistic** | **SE** | **z / chi-square** | **p** | **Corrected p** | **Result** |
| --- | --- | --- | --- | --- | --- | --- | --- |
| Main-effects LME | Time period omnibus | N/A | N/A | 161.59 | 1.25e-70 | 1.25e-66 | *** |
| Main-effects LME | 2 months post vs pre | 2.733 | 0.220 | 12.421 | 2.01e-35 | 2.01e-31 | *** |
| Main-effects LME | 6+ months post vs pre | -0.971 | 0.213 | -4.555 | 5.23e-06 | 0.0523 | n.s. |
| Hemisphere x time LME | Contralateral x 2 months post | -2.397 | 0.427 | -5.614 | 1.98e-08 | 1.98e-04 | *** |
| Hemisphere x time LME | Contralateral x 6+ months post | -3.228 | 0.413 | -7.805 | 5.93e-15 | 5.93e-11 | *** |

* Note: Below patient specific comparisons reported only for the above main significant omnibus effects.

| **Patient** | **Model / test** | **Effect / comparison** | **Estimate / statistic** | **SE** | **t / F** | **p** | **Corrected p** | **Result** |
| --- | --- | --- | --- | --- | --- | --- | --- | --- |
| P1 | Main-effects OLS | Time period omnibus | N/A | N/A | 114.07 | 8.49e-50 | 8.49e-46 | *** |
| P1 | Main-effects OLS | 2 months post vs pre | 5.4845 | 0.3714 | t = 14.7663 | 6.53e-49 | 6.53e-45 | *** |
| P1 | Main-effects OLS | 6+ months post vs pre | 2.8776 | 0.3777 | t = 7.6198 | 2.73e-14 | 2.73e-10 | *** |
| P1 | Hemisphere x time OLS | Contralateral x 2 months post | 2.4746 | 0.7423 | t = 3.3335 | 8.60e-4 | 1 | n.s. |
| P1 | Hemisphere x time OLS | Contralateral x 6+ months post | 0.2602 | 0.7548 | t = 0.3447 | 0.73 | 1 | n.s. |
| P2 | Main-effects OLS | Time period omnibus | N/A | N/A | 56.85 | 2.56e-25 | 2.56e-21 | *** |
| P2 | Main-effects OLS | 2 months post vs pre | 0.5648 | 0.3225 | t = 1.7513 | 0.08 | 1 | n.s. |
| P2 | Main-effects OLS | 6+ months post vs pre | -2.5511 | 0.2941 | t = -8.6740 | 4.62e-18 | 4.62e-14 | *** |
| P2 | Hemisphere x time OLS | Contralateral x 2 months post | -10.1513 | 0.6392 | t = -15.8818 | 2.60e-56 | 2.60e-52 | *** |
| P2 | Hemisphere x time OLS | Contralateral x 6+ months post | -5.6551 | 0.5829 | t = -9.7016 | 3.48e-22 | 3.48e-18 | *** |
| P3 | Main-effects OLS | Time period omnibus | N/A | N/A | 75.51 | 2.34e-33 | 2.34e-29 | *** |
| P3 | Main-effects OLS | 2 months post vs pre | 3.4321 | 0.4103 | t = 8.3655 | 6.51e-17 | 6.51e-13 | *** |
| P3 | Main-effects OLS | 6+ months post vs pre | -1.6119 | 0.4017 | t = -4.0123 | 6.04e-5 | 0.06 | n.s. |
| P3 | Hemisphere x time OLS | Contralateral x 2 months post | 1.2623 | 0.8201 | t = 1.5393 | 0.124 | 1 | n.s. |
| P3 | Hemisphere x time OLS | Contralateral x 6+ months post | -2.3296 | 0.8030 | t = -2.9009 | 0.004 | 1 | n.s. |

**(D) Planned post-operative auditory cortex (STG & HG) versus IFG time constant statistical testing results (see manuscript and Suppl. Fig. 7)**

Planned post-operative auditory cortex versus IFG time constant statistical testing results. Auditory cortex was defined as HG + STG; IFG was defined as IFGop + IFGtri. Positive estimates indicate longer time constants in IFG than auditory cortex; negative estimates show auditory cortex has shorter time constants than IFG. Corrected p-values are Bonferroni-adjusted across 20 planned tests (raw p x 20, capped at 1).

| **Time period** | **Contrast** | **Auditory mean** | **IFG mean** | **Estimate / beta** | **SE** | **z** | **p** | **Corrected p** | **Result** |
| --- | --- | --- | --- | --- | --- | --- | --- | --- | --- |
| Postoperative | 0.5*(IFGop + IFGtri) - 0.5*(HG + STG) | 18.0264 | 19.7169 | 1.6894 | 0.2720 | 6.2121 | 5.23e-10 | 1.05e-6 | *** |
| Postoperative | IFGop - 0.5*(HG + STG) | 18.0264 | 19.7169 | 0.6337 | 0.3331 | 1.9027 | 0.057 | 1 | n.s. |
| Postoperative | IFGtri - 0.5*(HG + STG) | 18.0264 | 19.7169 | 2.7451 | 0.3331 | 8.2419 | 1.69e-16 | 3.39e-12 | *** |
| 2 months post | 0.5*(IFGop + IFGtri) - 0.5*(HG + STG) | 18.3880 | 22.7323 | 4.3419 | 0.3917 | 11.0853 | 1.48e-28 | 2.96e-24 | *** |
| 2 months post | IFGop - 0.5*(HG + STG) | 18.3880 | 22.7323 | 3.2574 | 0.4797 | 6.7907 | 1.12e-11 | 2.23e-07 | *** |
| 2 months post | IFGtri - 0.5*(HG + STG) | 18.3880 | 22.7323 | 5.4264 | 0.4797 | 11.3124 | 1.14e-29 | 2.28e-25 | *** |
| 6+ months post | 0.5*(IFGop + IFGtri) - 0.5*(HG + STG) | 17.6972 | 16.9704 | -0.7268 | 0.3738 | -1.9446 | 0.052 | 1 | n.s. |
| 6+ months post | IFGop - 0.5*(HG + STG) | 17.6972 | 16.9704 | -1.7562 | 0.4578 | -3.8366 | 1.25e-4 | 1 | ** |
| 6+ months post | IFGtri - 0.5*(HG + STG) | 17.6972 | 16.9704 | 0.3026 | 0.4578 | 0.6611 | 0.509 | 1 | n.s. |

#### Supplementary Table 7. Time constant distribution Bowley distribution skewness test results

Planned patient-specific Bowley skewness was computed on log10(time constant). Positive delta Bowley indicates increased right-skew, consistent with fewer short time constants.

**(A) Time constant distribution skewness test results (manuscript Fig. 4H)**

| **Hemisphere** | **Patient** | **Comparison** | **Bowley A** | **Bowley B** | **Delta Bowley** | **95% CI** | **One-sided p** | **Two-sided p** | **Result** |
| --- | --- | --- | --- | --- | --- | --- | --- | --- | --- |
| Ipsilateral | P1 | Pre vs 2 months | -0.0285 | -0.0286 | -0.0001 | [-0.1146, 0.0859] | 0.4772 | 0.9980 | n.s. |
| Ipsilateral | P1 | Pre vs 6+ months | -0.0285 | 0.0691 | 0.0976 | [-0.0124, 0.1840] | 0.0212 | 0.0396 | * |
| Ipsilateral | P2 | Pre vs 2 months | -0.1635 | -0.0443 | 0.1192 | [0.0385, 0.2130] | 0.0019 | 0.0032 | ** |
| Ipsilateral | P2 | Pre vs 6+ months | -0.1635 | -0.1327 | 0.0308 | [-0.0467, 0.1124] | 0.1767 | 0.3640 | n.s. |
| Ipsilateral | P3 | Pre vs 2 months | -0.1056 | 0.2568 | 0.3624 | [0.2874, 0.4439] | 9.999e-05 | 9.999e-05 | *** |
| Ipsilateral | P3 | Pre vs 6+ months | -0.1056 | 0.0992 | 0.2048 | [0.1334, 0.2912] | 9.999e-05 | 9.999e-05 | *** |
